# Genetic pathways linking oxytocin-vasotocin hypothalamic subunit architecture with psychiatric and metabolic traits

**DOI:** 10.1101/2025.03.14.25323974

**Authors:** Alina I. Sartorius, Dennis van der Meer, Alexey Shadrin, Jaroslav Rokicki, Megan Campbell, Adriano Winterton, Ole A. Andreassen, Emanuel Schwarz, Terje Nærland, Lars T. Westlye, Daniel S. Quintana

**Author notes:** **For correspondence:** (DSQ).

## Abstract

The neuropeptides oxytocin and vasotocin are predominantly produced in the supraoptic and paraventricular nuclei of the anterior-inferior, anterior-superior and tubular-superior hypothalamic subunits. Evidence suggests that oxytocin and vasotocin signaling play a role in both physiology and behavior and that dysfunction of these signaling systems may contribute to the co-occurrence of metabolic and psychiatric conditions. The genetic pathways, however, that may underlie the connection between these physiological and behavioral traits are yet to be clearly delineated. We deployed bivariate mixture models and conjunctional FDR to estimate the global and local genetic overlap between three oxytocinergic-vasotocinergic hypothalamus subunits and ten psychiatric and metabolic traits related to oxytocin and vasotocin signaling. We show that these three subunits share moderate-to-extensive genetic overlap with the tested traits, therein stronger overlap with psychiatric than metabolic traits. We found most complete overlap between the anterior subunits and systolic blood pressure. We pinpoint 95 novel, unique loci associated across all subunit and trait combinations. The genes associated with these loci were enriched in gene sets linked to neuroimaging and neurodegeneration as well as metabolic markers, and were up-/down-regulated in tissues such as blood vessel and the liver. These findings help shed light on the genetic architecture of the hypothalamic subunits implicated in oxytocin and vasotocin, and selected traits, and provide new avenues for future research.

## Introduction

The neurotransmitter and hormone oxytocin has attracted increasing research attention due to its multifaceted role in health and well-being [1, 2]. Historically, oxytocin has been known for its functions in parturition [3] and lactation [4]. More recently, the neuropeptide has been popularized for its role in psychiatric conditions [5] and metabolic processes [6, 7], which frequently co-occur [8, 9, 10]. Recent evidence indicates that genetic variants in the oxytocin signaling pathway contribute to these co-occurrences and increased cardiovascular risk and genetic liability in certain psychiatric conditions [11]. Oxytocin, together with its paralog vasotocin [12], are among the most vital neuropeptides of the hypothalamus [13]. They are primarily synthesized in the hypothalamic paraventricular nuclei (PVN), in addition to the other major paraventricular peptide corticotropin-releasing hormone (CRH) [14], and they are exclusively synthesized in the hypothalamic supraoptic nuclei (SON) [15, 16] (***Figure 1***). Vasotocin is additionally synthesized on the suprachiasmatic nuclei (SCN) of the hypothalamus [17]. Together, the PVN and SON contain approximately 50,000 oxytocin neurons [18]. Oxytocin most likely emerged from the evolutionary ancestral oligopeptide vasotocin (also known as (arginine) vasopressin or antidiuretic hormone) by a gene duplication event [12, 19, 20]. Oxytocin and vasotocin are structurally almost identical [21, 22] and functionally very similar, too. Vasotocin was originally known for its physiological role in water homeostasis and the cardiovascular system [23, 24] as well as metabolism [25]. Recently, its contributions in behavioral functions (e.g., aggression, anxiety [26, 27]) have been increasingly explored alongside oxytocin. Moreover, oxytocin and vasotocin receptors can activate each other’s receptors, although affinities can vary. This facilitates cross-reactivity among the two peptide systems and a possible collaboration in the coordination of certain functions (e.g., social behavior) [28].

**Figure 1.**
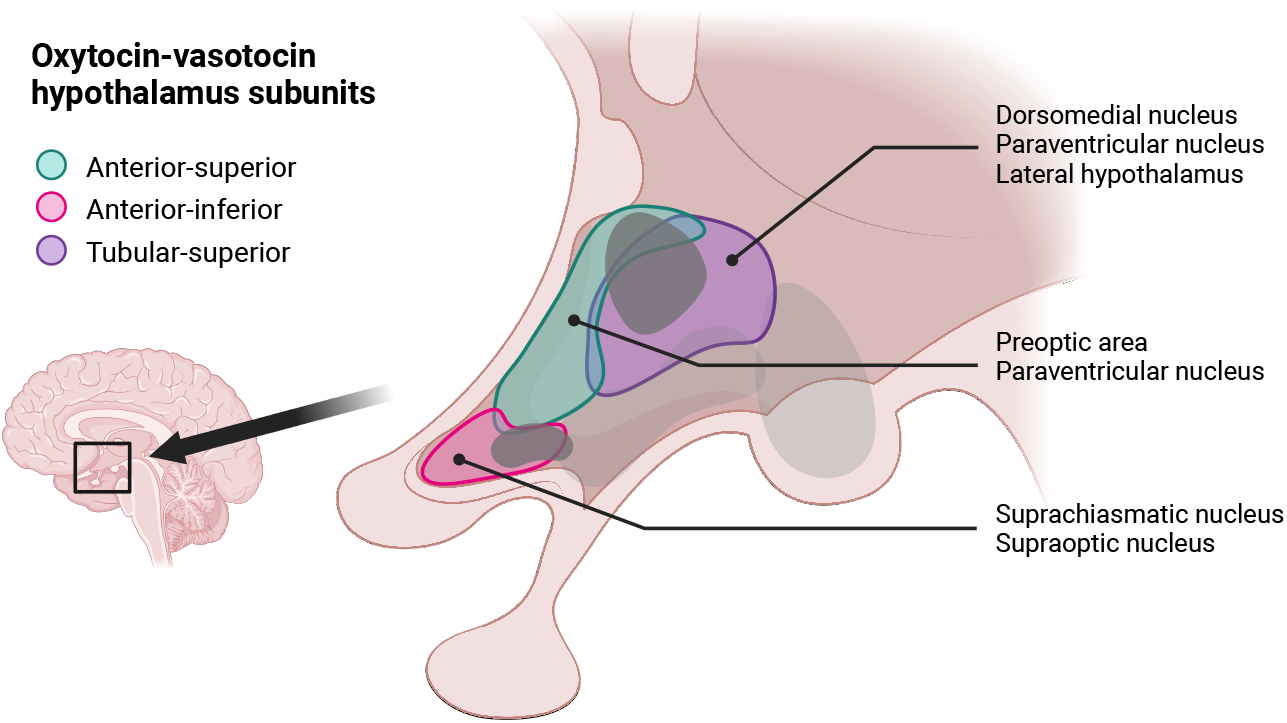
Anatomy of the human hypothalamus. Visible are the five subunits and the position of the two nuclei of interest (paraventricular nucleus, supraoptic nucleus) in the hypothalamus and its subunits. The aiHyp is depicted in magenta, asHyp in blue-green and tsHyp in purple. The tubular-inferior and posterior hypothalamic subunits are lightly grayed out since they were not analyzed here. The small upper dark-gray shaded area displays the paraventricular nucleus, the smaller lower dark-gray shaded area displays the supraoptic nucleus. Adapted from [60].

The hypothalamus is a subcortical structure located at the base of the brain that acts as a central hub for coordinating various complex physiological functions [29], such as eating behaviors [30, 31], sleep and wakefulness [32], body weight regulation [33, 34], cardiovascular regulation [35, 36], and endocrine systems [16, 37], and has been linked to psychiatric conditions [13]. There is evidence suggesting that the association between hypothalamic architecture and these conditions is mediated by oxytocinergic signaling. For instance, oxytocin and vasotocin may be linked to schizophrenia (SCZ) via an oxytocin-vasotocin degrading enzyme which is expressed in the PVN, SON and suprachiasmatic nucleus of the hypothalamus [38]. Moreover, stimulation of hypothalamic oxytocin neurons led to mania-/depression-like behavior in female mice [39], and the hypothalamus seems to regulate social cognition via oxytocin production and release, which may be relevant for psychiatric disorders and neurodevelopmental conditions characterized by difficulties with social behaviors ([40], but also see [13]). Regarding metabolic traits, knock-out rodent studies provide evidence for a link between the absence of paraventricular (but not supraoptic) released oxytocin and an increased body weight and food consumption, and between fewer paraventricular oxytocin neurons and higher body weight [41].

Despite the key role of these nuclei in orchestrating oxytocin signaling and these vital physiological and psychological processes, the genetic architecture of these nuclei, to a large degree, has not been well understood. One reason for this may be the small size of the nuclei, which has historically limited large-scale neuroimaging analysis to date. Only a few studies have investigated genetics in general in relation to the whole hypothalamus. Recently, a genome-wide association study (GWAS) [42] explored the general genetic architecture of different hypothalamic volumes, however, the study focused on the genetic correlation of whole hypothalamic volumes, and did not investigate the genetic overlap of selected hypothalamus subunits with psychiatric and metabolic traits. Subcortical structure volumes [43], as well as metabolic [44, 45] and psychiatric traits [46, 47], are moderately to highly heritable [48, 49]. Identifying the shared genetic architecture between the oxytocinergic-vasotocinergic hypothalamus subunits and psychiatric and metabolic traits — and subsequently functionally annotating them — can lead to a better understanding of disease mechanisms and diathesis. Computational advancements in brain imaging and segmentation methods together with the availability of large brain imaging and genetic data sets now allow for the characterization of hypothalamic substructure volumes at scale. This is an approach which our research group previously has shown to be instructive for other cerebral structures such as the thalamus [50], brain stem [51], basal ganglia [52], or cerebellum [53]. Research investigating the genetic overlap between two traits, such as brain substructures and psychiatric conditions, based on previously published GWAS data, has also proven useful in identifying novel genetic loci associated with various phenotypes such as SCZ and autism [54].

Here, we deploy a GWAS summary statistics based approach to quantify the genetic overlap between the three oxytocinergic-vasotocinergic hypothalamic subunits and both psychiatric and metabolic traits using causal mixture models (MiXeR [55]). MiXeR quantifies the genetic *overlap* (instead of correlation) between two traits, which facilitates the detection of mixed directions aof effect instead of only the same *or* opposite effect direction across the estimated shared genetic variants. This results in more nuanced and detailed results regarding the nature of overlap. We pinpoint pleiotropic loci that are jointly associated with a hypothalamus subunit and a trait using the conjunctional false discovery rate method (conjFDR [56]), which helps identify shared SNP-based biology between two traits. This type of approach is more powerful than a conventional, univariate GWAS and facilitates the discovery of pleiotropic loci that have not previously been described. Furthermore, we functionally annotate the novel loci and assess their expression across both the human brain and other tissues in the body [57, 58, 59]. Determining the crossover between genetic loci that jointly contribute to selected hypothalamus subunit structures and to psychiatric and metabolic phenotypes associated with the oxytocin/vasotocin system will better characterize the role of the oxytocin/vasotocin system and the pathophysiology in these co-occurring conditions.

## Results

### Hypothalamus subunit segmentation and genome-wide association study on oxytocinergic-vasotocinergic hypothalamus subunits

To facilitate the conduction of the core analyses — bivariate MiXeR and conjFDR —, the hypothalamus was segmented into its five subunits (see *Methods* section) and GWASs were performed on the three subunits of interest (anterior-inferior (aiHyp), anterior-superior (asHyp), tubular-superior (tsHyp)) to gain GWAS summary statistics for these hypothalamus volumes. Briefly, the GWAS for aiHyp revealed six significant LD-independent genomic loci associated with that subunit. We found 24 LD-independent loci to be significantly associated with the asHyp subunit. Lastly, 50 genomic loci were linked to tsHyp in the corresponding GWAS. Significant SNPs are reported in supplementary information 1 - 3, Manhattan and QQ plots are reported in supplementary information 4 - 6, GWAS summary statistics in supplementary information 7 - 9.

### Genetic overlap analysis using causal mixture models

We used the a priori generated GWAS summary statistics of the three hypothalamic subunits (aiHyp, asHyp, tsHyp) in combination with the genetic architecture of ten metabolic and psychiatric traits (autism, bipolar disorder (BD), SCZ, bone mineral density (BMD), body mass index (BMI), highdensity lipoprotein (HDL), low-density lipoprotein (LDL), systolic blood pressure (SBP), type II diabetes (T2D), and waist-to-hip ratio (WHR)) to assess the degree to which the traits genetically overlap. The three subunits were selected given their comprehensive coverage of the SON and PVN, which contain oxytocin and vasotocin neurons to a large degree, the SON exclusively [16, 61]. The PVN and SON, respectively, constitute large parts of the asHyp and aiHyp subunits. These subunits further include the preoptic area and suprachiasmatic nucleus, the latter also synthesizing vasotocin [17, 62, 63]. The tsHyp is mostly comprised of the posterior PVN, along with parts of the lateral hypothalamus and dorsomedial nucleus [62, 63]. For this analysis, we calculated bivariate causal mixture models using MiXeR [55]. The bivariate models build on univariate models conducted a priori, which gauge the number of variants linked to each phenotype individually. According to the generated heritability estimates and AIC values (supplementary information 10), the fit of the univariate MiXeR models for all thirteen traits individually was reliable, thus all traits were included in the subsequent bivariate MiXeR analyses [55].

The number of individual variants estimated for each trait are displayed in ***Figure 2***. Among the three hypothalamus subunits, with N_variants_ ≈ 2692 (SD ≈ 446, AIC = 11.76), most were estimated for the anterior-superior subunit, followed by N_variants_ ≈ 2414 (SD ≈ 495, AIC = 5.62) for the anterior-inferior and least for the tubular-superior subunit with N_variants_ ≈ 2124 (SD ≈ 210, AIC = 40.13). Among the psychiatric traits, autism was the most polygenic trait with N ≈ 12027 trait-influencing variants (SD ≈ 1284, AIC = 0.54), followed by SCZ and BD. Among the metabolic traits, BMI had the largest number of variants with an estimated N_variants_ ≈ 6552 (SD ≈ 632, AIC = 835.07), followed by WHR, SBP, BMD, T2D, HDL and LDL (see supplementary information 10 for all values).

**Figure 2.**
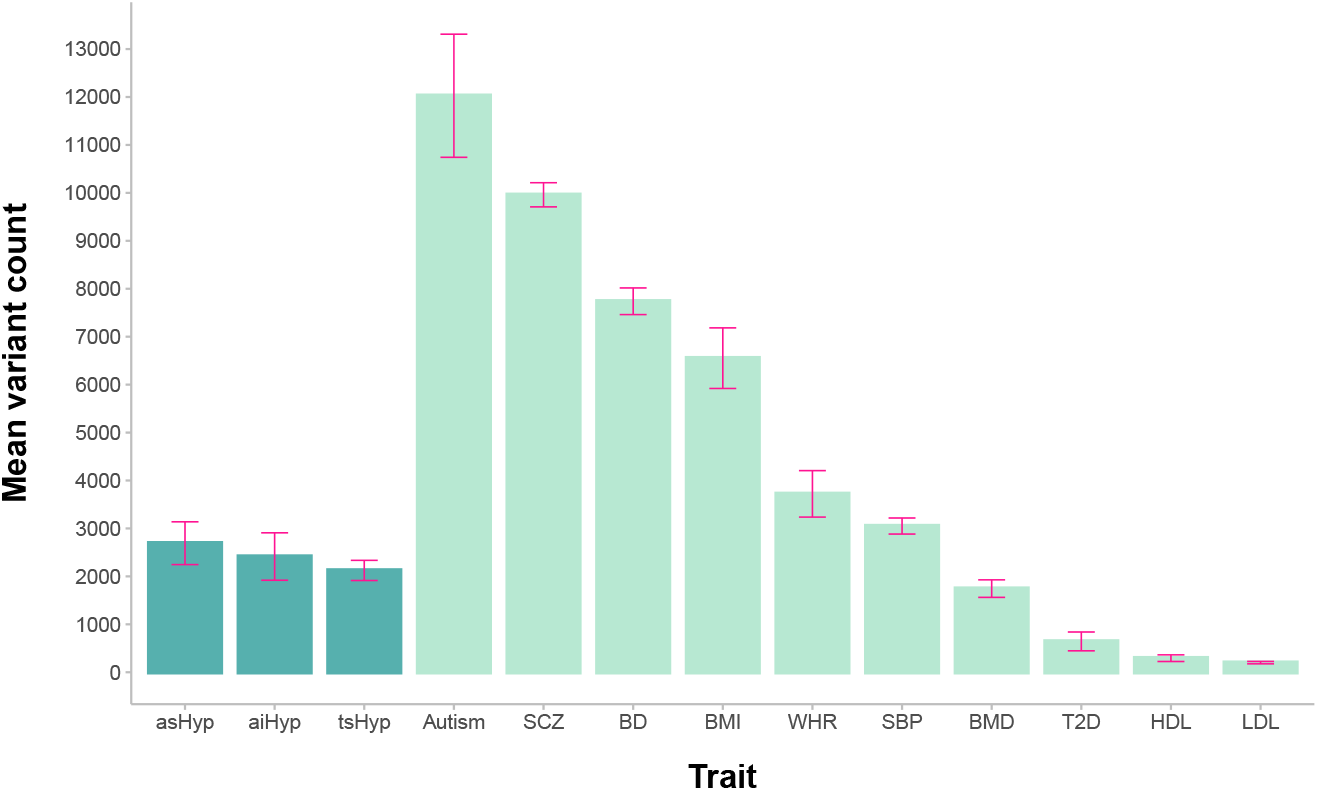
Estimated number of variants per trait. Mean variant counts (y-axis) resulting from the univariate MiXeR analyses for all 13 traits (x-axis). Estimates and corresponding standard deviations were obtained by averaging across 20 independent MiXeR runs. Darker blue-green columns refer to hypothalamus subunits, lighter blue-green columns refer to the psychiatric/metabolic traits, ordered from highest to lowest variant count. The standard deviations are displayed as magenta-colored whiskers. The data underlying this plot is available in supplementary information 30.

Continuing with the bivariate analyses, of the 30 MiXeR models (three hypothalamus subunits x ten psychiatric/metabolic traits, ***Figure 3***), seven models (aiHyp and autism/HDL, asHyp and autism/ HDL/LDL, tsHyp and HDL/LDL) had minimum and maximum AIC values *<* 0 (supplementary information 11) and/or a non-credible log-likelihood distribution, and were thus discarded. The remaining 23 reliable bivariate models showed varying degrees of genetic overlap between the three hypothalamus subunits and eight psychiatric/metabolic traits.

**Figure 3.**
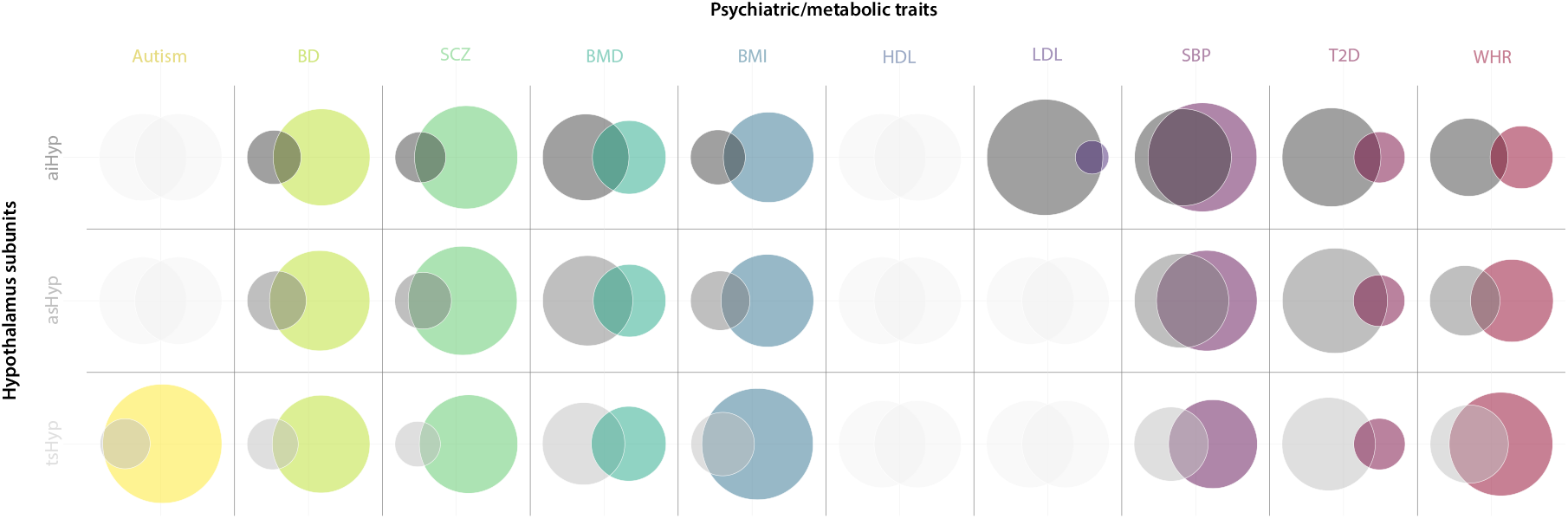
Genetic overlap of the bivariate MiXeR analyses. The Venn diagrams visualize the degree of genetic overlap between two traits. *relation to* the other circle, the more/fewer variants have been found to influence a particular trait. The more enetic variants two traits share and the less trait-unique variants there are, indicating a higher degree of genetic diagram for the asHyp subunit and WHR shows two circles comparable in size, with WHR being slightly larger, ariants influencing each trait and WHR being more polygenic (approximately 1842 and 2871 SNPs), and a _red_ ≈ 851). Missing Venn diagrams due to unreliable model fit are indicated with white-grey dummy circles. The re displayed in rows one to three in dark, medium and light grey, respectively; the ten metabolic and psychiatric umns one to ten in yellow, lime green, green, teal, teal blue, dark blue (HDL, not displayed due to unreliable rgundy and dark red. Exact numbers of unique and shared variants for each comparison as well as the data producing the Venn diagrams is available in supplementary information 12.

Generally, the genetic overlap varied and ranged from small to extensive (1). The most complete overlap was found between the two anterior subunits and SBP (aiHyp: N ≈ 2046 shared variants, 369 and 1005 unique; asHyp: N ≈ 1929 shared variants, 764 and 1122 unique), which is also reflected in the minimum and maximum AIC values, where the max AICs are smaller and the min AICs are considerably larger. This indicates a strong overlap and only few variants that are unique to both individual traits. The fraction concordance rates (47.48% and 49.13% for aiHyp and asHyp) point to an equal number of the shared variants having the same direction and an opposite direction of effect. Furthermore, almost all variants from the tsHyp subunit were covered by the variants estimated for autism (97.34%), however, autism only shared 17.19% of its variants with tsHyp. A similar, less extreme pattern could be observed for tsHyp with BMI (84.97% and 27.55% variants shared), asHyp with SCZ (77.55% and 20.96% variants shared), and reversed for LDL with aiHyp (86.2% and 7.16% variants shared). The least overlap was observed between aiHyp and WHR (N ≈ 438 shared variants, 1976 and 3284 unique), with a negative min AIC value and a positive AIC value, indicating the presence of non-complete overlap with more trait-specific variants.

The overlap patterns from the Venn diagrams of the two anterior subunits were noticeably uniform across psychiatric and metabolic traits with a consistently comparable degree of overlap. There were some differences in the pattern of overlap between the two anterior subunits and the tubular subunit, particularly in the traits BMI, SBP and WHR. The two anterior subunits generally overlapped less with BMI and WHR and more with SBP, while the opposite was the case for the tubular-superior subunit. For instance in BMI, aiHyp and asHyp shared 31.77% and 41.4%, respectively, of the variants with the metabolic trait, while tsHyp shared 84.97% (more than double). For SBP on the other hand, the variants estimated for tsHyp overlapped only to 44.86%, but to 84.73% and 71.62%, respectively, for aiHyp and asHyp. In the remaining traits (BD, SCZ, BMD, T2D), no clear grouping of overlap between anterior versus tubular subunits was discernible. Particularly for BMD and T2D did all three subunits show similar patterns and degrees of overlap (***Figure 3***).

All three hypothalamus subunits, averaged separately across metabolic and psychiatric traits, respectively, shared relatively more of their variants with the psychiatric traits (aiHyp: 52.58%, asHyp: 68.11%, tsHyp: 61.05%) than with the metabolic traits (aiHyp: 30.63%, asHyp: 38.06%, tsHyp: 48.45%), most pronounced in the anterior-superior subunit. The fraction of concordance on the other hand was slightly higher with metabolic traits for all subunits. aiHyp, asHyp and tsHyp shared ∅ 37.40%, ∅ 41.21% and ∅ 46.13% concordant variants across psychiatric traits and ∅ 53.97%, ∅ 46.09% and ∅ 47.82% across metabolic traits.

Among the hypothalamic subunits, aiHyp shared the biggest fraction of trait-influencing variants with the metabolic trait SBP, followed by BMI, BMD, WHR, T2D and LDL. Among the two psychiatric traits, SCZ had the bigger share over BD (table 1). asHyp genetically overlapped the most with SBP among the metabolic traits, followed by BMI, WHR, BMD and T2D, and among the psychiatric traits the most with SCZ, followed by BD (table 1). Lastly, tsHyp shared most genetic variants with the metabolic trait BMI, followed by WHR, SBP, BMD, and T2D. Regarding psychiatric traits, tsHyp had the largest genetic overlap with autism, followed by BD and SCZ (table 1) (refer to supplementary information 11 and 12 for detailed MiXeR results and supplementary information 13 for genetic correlation).

**Table 1.**
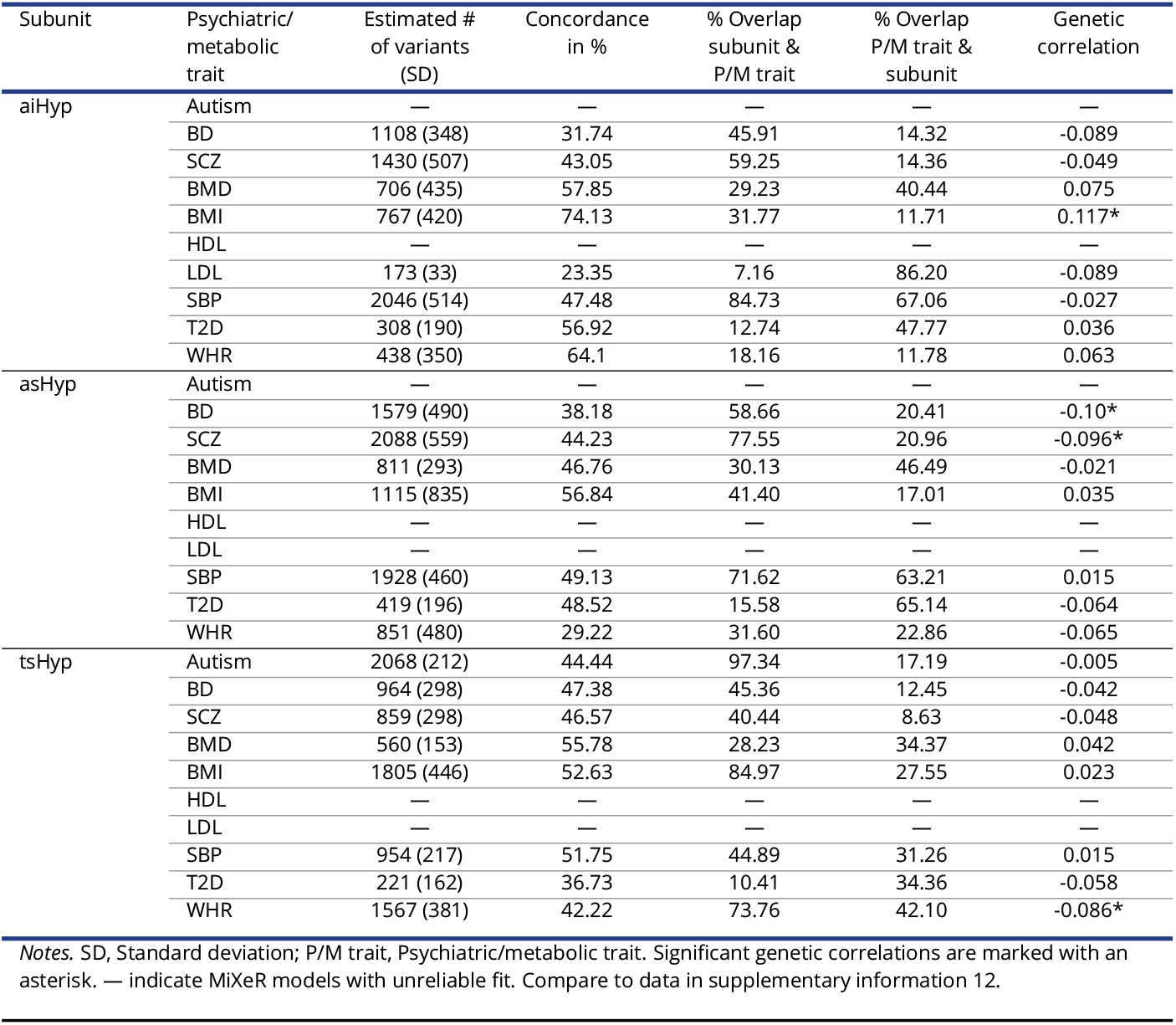
Overview of genetic overlap from MiXeR and genetic correlation from LDSC regression.

### Conjunctional false discovery rate for detection of shared genetic loci

The conjFDR method complements a bird’s-eye overview of the shared genetic architecture assessed by MiXeR by identifying specific loci shared between two traits. ConjFDR analyses revealed 107 significant shared loci across all subunit and trait combinations. Of those, 95 were unique. All models including BMD were removed due to UKB sample overlap with the hypothalamus subunits, and aiHyp and autism/HDL, asHyp and autism/HDL/LDL, and tsHyp and HDL/LDL were not considered due to unreliable model fit from previous MiXeR analyses (see *Methods* section). Nine loci were repeatedly identified in different subunit and trait combinations. For the hypothalamus subunits specifically, aiHyp analysis revealed 30 variants (26 unique) jointly associated with the different metabolic and psychiatric traits, of which three SNPs were shared with BD, seven shared SNPs with SCZ, four shared with BMI, four shared SNPs with LDL, eight shared SNPs with SBP, two shared SNPs with T2D, and two shared SNPs with WHR. For the hypothalamus subunit asHyp, the tests yielded in total 33 variants (31 unique) which were linked to both the subunit architecture and a psychiatric/metabolic trait. Specifically, three SNPs were shared with the trait BD, 17 shared SNPs with SCZ, five shared SNPs with BMI, five shared SNPs with SBP, one shared SNP with T2D, and two shared SNPs with WHR (***Figure 4***). Lastly, for the subunit tsHyp, the conjFDR analysis revealed in total 44 variants (43 unique) of which four SNPs were shared with autism, one SNP was shared with BD, 14 shared SNPs with SCZ, nine shared SNPs with BMI, nine SNPs with SBP, one SNP with T2D, and six shared SNPs with WHR (see supplementary information 14 for full list of rsIDs, *p*-values and further chromosomal information).

**Figure 4.**
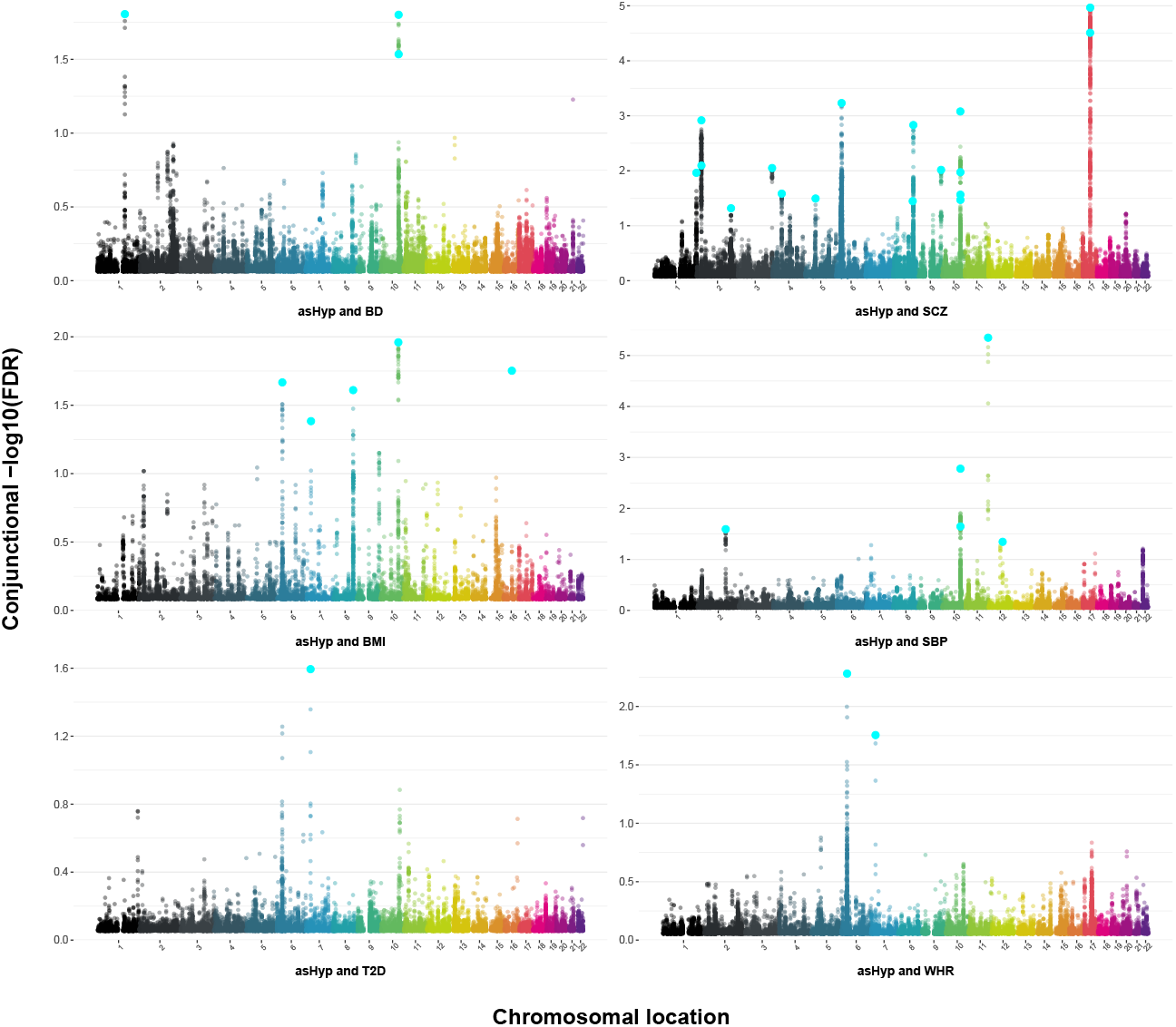
Manhattan plots for asHyp and the six traits that passed AIC/log-likelihood without UKB sample overlap. Distribution of the conjFDR values (y-axis) for each tested variant across the human genome in six independent Manhattan plots. One dot represents one variant, with the variants/loci passing the significance threshold being highlighted in bright cyan in each plot. Chromosomes (x-axis) are displayed in different colors. For instance, in the Manhattan plot for the subunit asHyp and T2D (bottom outer left plot), one bright cyan dot (i.e., locus) is visible that was statistically significant in this comparison. The raw data underlying this plot is presented in supplementary information 31. Manhattan plots for the other two subunits are presented in supplementary information 32 (raw data in supplementary information 33) and supplementary information 34 (raw data in supplementary information 35).

### Annotation of cross-trait significant loci

We proceeded with the annotation of the unique loci per subunit and psychiatric traits, and per subunit and metabolic traits in order to determine their functional relevance. Using the ‘Variant to Gene’ (V2G) score integrated into Open Target Genetics [57, 58], we searched for all relevant genes surrounding a variant of interest. After the exclusion of six variants due to missing data, the removal of duplicate genes within a gene set of subunit and psychiatric or metabolic traits, and the exclusion of genes that did not meet the cut-off values (see *Methods* section for filter and cut-off information), this led to the identification of 26 unique genes linked to conjFDR variants identified in aiHyp with psychiatric traits and 125 unique genes with metabolic traits, 182 unique genes linked to conjFDR variants identified in asHyp with psychiatric traits and again 67 unique genes with metabolic traits, and 70 unique genes linked to conjFDR variants identified in tsHyp with psychiatric traits and 151 unique genes with metabolic traits (for the raw lists including duplicates, see supplementary information 15). Subsequently, to highlight potentially relevant biological mechanisms genes identified for each hypothalamus subunit and psychiatric/metabolic trait groups (six gene sets in total) were analyzed using FUMA (“Functional Mapping and Annotation of Genome-Wide Associations Studies”) [59] GENE2FUNC.

We were particularly interested in the tissue specificity annotation and gene set enrichment. For the genes from the aiHyp and psychiatric traits gene sets, the tissue specificity annotation showed that across 30 tissue types in the human body, the gene set was significantly down-regulated in the esophagus and salivary gland with seven (*p* = 4.01e-2) and eight genes (*p* = 1.55e-2). The gene set was not significantly up-regulated in any tissue. The genes from the gene set of the conjFDR for aiHyp and metabolic traits were significantly down-regulated in the spleen with 14 genes (*p* = 3.23e-2), and not significantly up-regulated in any of the 30 tested tissues. Of the gene set from the conjFDR for asHyp and metabolic traits, eight genes were up-regulated in blood vessel (*p* = 3.50e-3, ***Figure 5***), and none were significantly down-regulated in any tissue. The genes annotated to the conjFDR for tsHyp and psychiatric traits were significantly down-regulated in the liver with 36 genes (*p* = 4.03e-2; all *p*-values Bonferroni corrected); the gene set was not significantly up-regulated in any tissue. The two remaining gene sets (asHyp and psychiatric traits, tsHyp and metabolic traits) were not significantly up- or down-regulated in any of the 30 tissue types (view supplementary information 16 - 21 for detailed result statistics).

**Figure 5.**
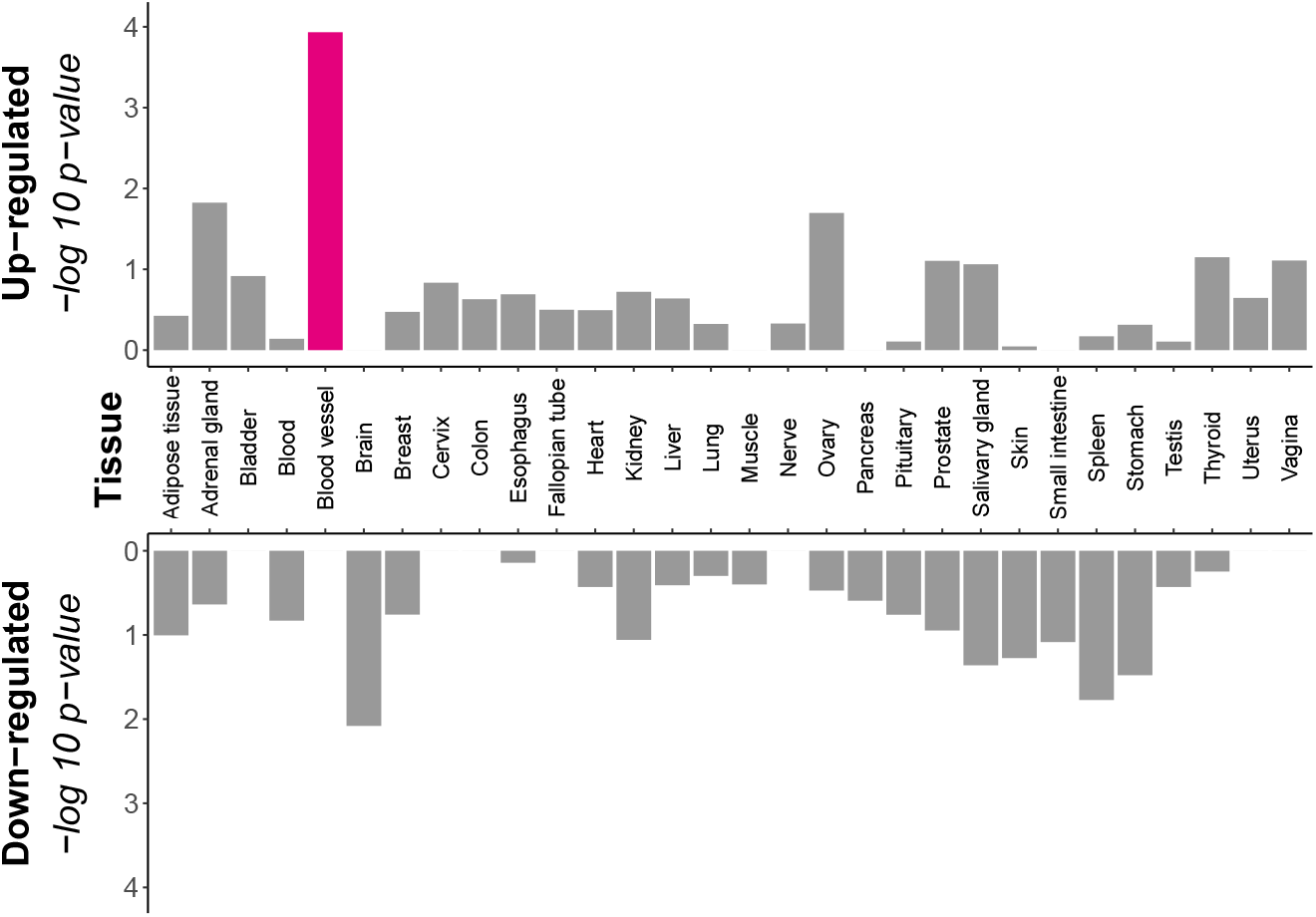
Tissue specific up- and down-regulation of genes associated with the asHyp and metabolic traits conjFDR analysis. The 67 credible, filtered genes that were linked to the variants produced by the conjFDR analysis for the asHyp subunit with the four metabolic traits BMI, SBP, T2D, and WHR display varying degrees of enrichment and reduction across the 30 human tissues tested with FUMA. Columns for tissues that were significantly up-regulated are colored in magenta (i.e., blood vessel), non-significant tissues are colored grey. Up-regulation is displayed at the top, down-regulation at the bottom of the plot. *p*-values shown on the y-axes were log-transformed for visualization purposes, tissues on the x-axis are in alphabetical order. The raw data underlying this plot is presented in supplementary information 36. Figures and plot data for the remaining five gene sets are provided in supplementary information 37 - 46.

GWAS gene set enrichment assessments revealed a significant overlap between the aiHyp and psychiatric traits gene set (n_genes_ = 26) and 67 GWAS catalog gene sets. The top five most significant enriched GWAS gene sets were “Plasma omega-3 polyunsaturated fatty acid levels (docosapentaenoic acid)” with six overlapping genes (*p* = 2.85e-11), “Plasma omega-6 polyunsaturated fatty acid levels (arachidonic acid)” with six overlapping genes (*p* = 7.71e-11), “Autism spectrum disorder or schizophrenia” with 11 overlapping genes (*p* = 9.03e-11), “Waist-to-hip ratio adjusted for BMI (age >50)” with nine overlapping genes (*p* = 2.71e-10), and “Brain morphology (MOSTest)” with 14 overlapping genes (*p* = 4.81e-10; all *p*-values FDR-corrected for multiple testing, see supplementary information 22). For the 125 genes associated with the aiHyp and metabolic traits subset, we found overlap with 83 GWAS gene sets, with the top five “Plasma omega-6 polyunsaturated fatty acid levels (arachidonic acid)” with nine overlapping genes (*p* = 2.41e-15), “Plasma omega-3 polyunsaturated fatty acid levels (docosapentaenoic acid)” with eight overlapping genes (*p* = 3.50e-14), “Plasma omega-6 polyunsaturated fatty acid levels (linoleic acid)” with eight overlapping genes (*p* = 7.28e-13), “Waist-to-hip ratio adjusted for BMI” with 20 overlapping genes (*p* = 1.50e-12), and “Plasma omega-6 polyunsaturated fatty acid levels (dihomo-gamma-linolenic acid)” with seven overlapping genes (*p* = 1.00e-11; all *p*-values FDR-corrected for multiple testing, see supplementary information 23).

The 182 genes linked to the asHyp and psychiatric traits gene subset were significantly enriched in 54 GWAS gene sets, the top five being “Brain morphology (MOSTest)” with 53 overlapping genes (*p* = 5.06e-29), “Cortical surface area (MOSTest)” with 29 overlapping genes (*p* = 2.53e-18), “Subcortical volume (MOSTest)” with 29 overlapping genes (*p* = 2.39e-15), “Brain morphology (min-P)” with 24 overlapping genes (*p* = 1.43e-14), and “Cortical thickness (MOSTest)” with 20 overlapping genes (*p* = 1.81e-14; all *p*-values FDR-corrected for multiple testing, see supplementary information 24). For the 67 genes associated with the asHyp and metabolic traits subset, we found a significant enrichment in 33 GWAS gene sets, the top five being “Mean arterial pressure” with six overlapping genes (*p* = 7.47e-7), “Systolic blood pressure” with nine overlapping genes (*p* = 9.53e-7), “Brain morphology (MOSTest)” with ten overlapping genes (*p* = 1.06e-6), “Blood pressure” with five overlapping genes (*p* = 2.79-6), and “Waist-to-hip ratio adjusted for BMI (age >50)” with six overlapping genes (*p* = 2.91e-6; all *p*-values FDR-corrected for multiple testing, see supplementary information 25).

Lastly, the 70 genes linked to the tsHyp and psychiatric traits gene set were statistically significantly enriched in 43 GWAS gene sets. Here, the top five gene sets were “Autism spectrum disorder or schizophrenia” with 20 overlapping genes (*p* = 6.76e-17), “Schizophrenia” with 22 overlapping genes (*p* = 4.47e-15), “Brain morphology (MOSTest)” with 26 overlapping genes (*p* = 8.40e-15), “Parkinson’s disease” with ten overlapping genes (*p* = 2.11e-8), and “Response to cognitive-behavioural therapy in major depressive disorder” with six overlapping genes (*p* = 3.87e-7; all *p*-values FDR-corrected for multiple testing, see supplementary information 26). The 151 genes associated with the tsHyp and metabolic traits gene set were significantly enriched in 111 GWAS sets. The top five gene sets here were “Carotid intima media thickness (maximum)” with eight genes overlapping (*p* = 1.56e-12), “Cerebrospinal fluid t-tau levels in mild cognitive impairment” with eight overlapping genes (*p* = 8.46e-12), “Cerebrospinal AB1-42 levels in mild cognitive impairment” with eight genes overlapping (*p* = 1.46e-11), “Cerebrospinal AB1-42 levels in Alzheimer’s disease dementia” with eight overlapping genes (*p* = 2.53e-11), and “Logical memory (immediate recall)” with eight genes overlapping (*p* = 8.58e-11; *p*-value FDR-corrected for multiple testing, see supplementary information 27).

## Discussion

Here we systematically explored the shared genetic architecture underlying selected psychiatric/metabolic traits and the structure of oxytocinergic-vasotocinergic hypothalamic subunits. Oxytocin’s role in metabolism and different psychiatric phenotypes has been previously described [11, 64], however, the pathways underlying this connection remain mostly unclear. The hypothalamic paraventricular (PVN) and supraoptic (SON) nuclei largely [61] or exclusively [16] contain oxytocin and vasotocin neurons and have been implicated in several metabolic and cardiovascular phenotypes as well as mental health [6]. Metabolic and psychiatric traits, as well as subcortical brain region morphology, are typically characterized by significant heritability estimates [43, 44, 45, 46, 47]. Thus, investigating the variants and genes that these two trait subsets (hypothalamus subunits and psychiatric/metabolic traits) may have in common can be instructive in uncovering aspects of the genetic pathways linking the oxytocin-vasotocin system to metabolism and psychiatric conditions. We found sizable differences in polygenicity within traits and varying degrees of genetic overlap between traits. These findings were extended by conjFDR analyses and subsequent functional annotation, which identified significant individual SNPs that were linked to genes, which were assessed regarding their differential expression in the human body and their enrichment in different GWAS.

Univariate MiXeR analyses revealed that the 13 initially included traits (hypothalamus subunits aiHyp, asHyp, tsHyp, and psychiatric/metabolic traits autism, BD, BMD, BMI, HDL, LDL, SBP, SCZ, T2D, WHR) are associated with a wide range of trait polygenicity, ranging from 200 loci estimated for LDL being the least polygenic, to 12027 loci for autism having the most polygenic nature. The top three polygenic traits in this analysis were the three psychiatric traits: autism, SCZ, and BD. Both anterior hypothalamic subunits were more polygenic than the tubular-superior subunit. Complex and heterogeneous phenotypes have been shown to be highly polygenic [65] and to be influenced by large numbers of common variants with small effects [66]. Accordingly, psychiatric conditions have been previously shown to have a polygenic nature [67]. A key defining attribute of autism, for instance, is indeed the wide, heterogeneous spectrum of features [68, 69]. Similarly, SCZ and

BD are both characterized by a wide spectrum of symptoms (e.g., ‘positive’ symptoms including hallucinations and delusions and ‘negative’ symptoms resembling depression, but also cognitive dysfunction [70, 71]). Apart from this, negative selection has been discussed as a factor explaining polygenicity in biological complexity [65, 72].

Using these variants, bivariate causal mixture models for the 23 MiXeR comparisons with reliable model fit revealed that, on average, the hypothalamus subunits shared more of their variants with the psychiatric traits than with the metabolic traits, ranging from 12.60% difference between psychiatric and metabolic in tsHyp, over 21.95% in aiHyp, to 30.04% in asHyp. Given these moderate to large differences, the genetic pathways that may connect the subunits with the psychiatric traits can be considered more polygenic than with the metabolic traits, which in part could be explained by the higher baseline polygenicity of the psychiatric traits (compare to ***Figure 2***). We found the closest to complete overlap between any hypothalamus subunit and any psychiatric/metabolic trait for the anterior-inferior subunit and SBP. This points to a very similar variant-based genetic architecture of the two traits and may be indicative of a common underlying genetic pathway. In the majority of the comparisons, the number of shared variants with the same direction of effect was similar to the number of shared variants with the opposite direction of effect. This is also reflected in genetic correlations that were often close to zero (1. This points to the ‘ability’ of a common genetic system or blueprint, which consists of the same variants, to exert contrary effects in two traits depending on the functionality of the trait, highlighting the versatility and complexity of these genetic systems. We further found considerable overlap of 97% for tsHyp variants with autism variants, meaning that almost all of the variants estimated for the tubular-superior subunit were shared with autism variants. Importantly, at the same time, only 17% of the variants estimated for autism overlapped with tsHyp, while the overall overlap between this subunit and psychiatric trait was moderate (compare to 1). This pattern is most likely again explained by the high polygenicity (or even omnigenicity [73]) of autism and the comparably markedly lower polygenicity of the tubular-superior subunit, which may lead to one trait appearing to be “inside” of another trait (see ***Figure 3***). Notably, aiHyp shared the majority of variants with autism, too, however, this model did not pass quality control checks. Overlap patterns between the two anterior subunits and the tubular subunit were similar across traits except for BMI, SBP and WHR (***Figure 3***). The tsHyp subunit showed a more extensive overlap with the two measures of body composition compared to the anterior subunits, and vice versa for blood pressure, potentially pointing to a specific link between the two anterior subunits and blood pressure, and between the tubular-superior subunit and body composition. The two psychiatric traits BD and SCZ show comparable overlap patterns with each respective hypothalamus subunit. Previous work has reported a near identical genetic architecture between BD and SCZ [74]. Thus, it appears reasonable that the two traits would behave similarly to the three subunits and display similar genetic overlap. Lastly, genetic correlations were non-significant for the majority (19 out of 23) of trait pairs. The presence of genetic overlap in the absence of significant genetic correlation may indicate that the genetic overlap goes beyond correlation, suggesting a complex “non-uniform” interaction of the different phenotypes.

Deploying a conjunctional FDR (conjFDR) approach, we identified specific loci that are shared between a hypothalamus subunit and a trait. Across all subunits and traits, we pinpointed 95 unique SNPs. In line with the results from the bivariate MiXeR models, where tsHyp had most extensive most genetic overlap across traits, followed by asHyp and aiHyp last, tsHyp also shared most loci across traits here (43 unique SNPs), followed by asHyp (31 unique SNPs) and aiHyp (26 unique SNPs). Across subunits, most SNPs were identified in association with SCZ, followed by SBP and BMI. Interestingly, BD was among the traits with least joint SNPs, although it has been shown to have an almost identical set of loci as SCZ [74]. Considering that, in conjFDR, as compared to MiXeR, the yield of loci primarily depends on the power of the underlying GWAS summary statistics, this could be due to the SCZ GWAS being more powerful compared to the BD GWAS.

Subsequently, we annotated the loci from the conjFDR analyses with meaningful, functionally relevant genes. Grouped into six subsets by the three subunits (aiHyp, asHyp, tsHyp) with either psychiatric (autism, BD, SCZ) or metabolic traits (BMD, BMI, HDL, LDL, SBP, T2D, WHR), we linked 26, 125, 182, 67, 70 and 151 unique genes to the SNPs in the six subsets. Functional annotation of the gene sets using FUMA revealed up- or down-regulation in four out of the six submitted gene sets in some tissue out of 30 tissues across the human body, down-regulation being more common. The gene set associated with the asHyp subunit and metabolic traits was the only one that was significantly up-regulated, specifically, in blood vessel tissue. This may be in line with the highly significant enrichment of this gene set in different GWAS gene sets regarding measures of blood pressure (e.g., arterial pressure, hypertension, pulse pressure) and cardiovascular health (e.g., coronary heart disease, carotid plaque burden, myocardial infarction) that we found in the FUMA gene set enrichment analysis. Further, we found down-regulation for both gene sets from the conjFDR analysis of the aiHyp subunit with psychiatric (in the esophagus, salivary gland) and metabolic traits (in the spleen). The down-regulation of genes (e.g., *FADS2, FADS3, MYRF, BEST1*) of the former gene set in the esophagus and salivary gland could depict an association with metabolism and diabetes mellitus [75] (as shown in rodents), which would also be reflected in the highly significant enrichment of those genes in GWAS catalog gene sets related to fatty acid levels, metabolites, or body composition. On the other hand, some of the genes down-regulated in these tissues (e.g., *CYP17A1, INA, CACNB2*) we found to be enriched in GWAS gene sets related to cardiovascular and psychiatric traits. We found similar results for the latter gene set (conjFDR analysis for aiHyp and metabolic traits), which was down-regulated in the spleen and enriched in GWAS gene sets related to fatty acid levels, metabolism, cardiovascular health and psychiatric traits. Furthermore, some of the genes from the gene set derived from the conjFDR analysis for tsHyp and psychiatric traits were down-regulated in the liver, which is a central metabolic but also endocrine organ in the human body [76], again possibly reflecting the gene’s role in metabolism but also the endocrine system. This, however, was not particularly mirrored in the enrichment analysis, where a different subset of genes were enriched in gene sets associated with a range of traits (e.g., psychiatric phenotypes, cognitive ability, body composition, cancer, fibrosis).

The two remaining gene sets — from the conjFDR for asHyp and psychiatric traits and for tsHyp and metabolic traits — that were not up- or down-regulated in any tissue, did, nevertheless, show distinct GWAS gene set enrichment patterns. For the genes from the asHyp and psychiatric traits conjFDR we observed a particular enrichment in GWAS gene sets featuring measures of cortical morphology and anatomy. A subset of genes from the tsHyp and metabolic traits conjFDR gene set was distinctly enriched in GWAS gene sets related to cognitive performance and neurodegeneration (in addition to other, more non-specific gene sets which also appeared in all results for the other gene sets). These genes have been previously linked to Alzheimer’s disease (e.g., *APOE* [77], *BCAM* [78], *TOMM40* [79], *APOC1* [79], *CBLC* [78]). Taken together, all six gene sets were enriched in a varied range of GWAS gene sets (e.g., fatty acids, brain morphology, cognition, male-pattern baldness, handedness, Parkinson’s disease). These varied outcomes could be due to other hypothalamic nuclei (e.g., preoptic area, lateral hypothalamus) being part of the analyzed subunits [62, 63] (discussed in detail below), which may explain why the gene sets were enriched in somewhat heterogeneous GWAS gene sets.

There are some limitations to this study that are worth noting. Due to the nature of the conjFDR analyses, the samples which the GWAS summary statistics of the different phenotypes are based on should not genetically overlap [55], since any sample overlap may introduce some inflation of the results. Given that the GWAS summary statistics for the three hypothalamus subunits are derived from UK Biobank (UKB), for all other traits, the GWAS summary statistics could not be taken from genetic association data sourced from the UKB. This has proven to be especially intricate for the trait BMD (refer to the *Methods* section). To our knowledge, no summary statistics for the trait BMD from a large GWAS were available without drawing on UKB data. Thus, BMD was excluded from all conjFDR analyses. This limitation will persist until prospective large GWAS on BMD excluding UKB data are published.

The neuronal population in the three subunits we chose to represent the hypothalamic oxytocinvasotocin neural network are not limited to oxytocin and vasotocin neurons. The PVN, for instance, also produces the corticotropin-releasing hormone (CRH [14]) and to a lesser extent the thyrotropin-releasing hormone (TRH [80]. The three subunits harbor additional nuclei such as the suprachiasmatic nucleus (which contains some vasotocin neurons [17], too) or the dorsomedial nucleus (mainly a ‘distribution point’ over a peptide synthesis site and an important regulatory center for processing signals from hormones synthesized elsewhere in the hypothalamus (e.g., acetylcholine [81], glutamate [82])). The current standards of imaging and automated segmentation tools facilitate the segmentation of brain structures as small as the hypothalamus into five smaller sub-structures, however, the available tools reach their limits when it comes to finer parcellation down to the nucleus level. Until imaging and segmentation techniques advance further, our approach deployed here is the closest approximation currently available to isolate oxytocinergic/vasotocinergic hypothalamus volumes in a large dataset.

In studies investigating the genetic correlation between two traits, cross-trait assortative mating must be considered as an alternative mechanism besides or instead of pleiotropy. Pleiotropy is the notion that one locus influences two or more phenotypes, and is commonly portrayed as the descriptive mechanism underlying observed genetic correlation. Cross-trait assortative mating refers to cross-correlation across phenotypes among mating partners [83]. There has been evidence to suggest that cross-trait assortative mating can account for a considerable proportion of variation in genetic correlation [83], calling into question the basic assumption that individuals mate at random as well as the robustness of genetic correlation estimates and their derived interpretations regarding genetic mechanisms. In this study, although we focused on genetic overlap rather than genetic correlation, the putative effect of cross-trait assortative mating on our analyses and interpretation of the results should not be overlooked.

A final limitation is that since all GWAS summary statistics included were based on European/white cohorts it follows that the results from all analyses is limited to individuals of European/white descent, and that the results might not be generalizable to different populations.

This study advances our understanding of the genetic underpinnings of the hypothalamus subunits in relation to selected metabolic and psychiatric traits. We applied a novel approach by investigating the genetic overlap between specific subunits — instead of whole structure volumes — of the hypothalamus that orchestrate the oxytocin and vasotocin production and psychiatric/metabolic traits. This subunit-level analysis allowed for a more targeted investigation of the oxytocin-vasotocin system. Prior research has found that the oxytocin genetic pathway is associated with different metabolic and psychiatric traits [11] and that vasotocin is involved in similar traits [23, 24, 25, 26, 27]. Our research is consistent with this and extends these findings by quantifying the genetic overlap and pinpointing the SNPs that may constitute this genetic pathway, considering the specific oxytocin/vasotocin synthesis sites in the hypothalamus. All three subunits shared more genetic overlap with psychiatric traits than with metabolic traits, and we identified 95 novel, unique SNPs associated jointly with the three subunits and psychiatric/metabolic traits. We functionally annotated the detected SNPs and showed that some of the associated genes are up-regulated in blood vessel tissue and down-regulated among others in the esophagus and liver. The genes were enriched in diverse traits depending on the underlying conjFDR analysis, and in traits beyond, such as cognitive decline, brain morphology and further psychiatric and neurological conditions. These findings emphasize the polygenic architecture of the oxytocin/vasotocin hypothalamic subunits and health-relevant traits, such as schizophrenia and blood lipid levels. With the identified SNPs, we provide new potential avenues for future research and help shed light on the intricate role the oxytocinergic-vasotocinergic hypothalamus systems in health and well-being.

## Methods

If not stated otherwise, the statistical software R (versions 4.3.1 and 4.4.1 [84]), RStudio (versions 2023.06.1+524 and 2023.09.1+494 [85]), FreeSurfer (version 7.4.1 [62]), REGENIE (version 3.6 [86]), MiXeR (version 1.3 [55]), LDSC (version version 1.0.1 [87, 88, 89]) and conjFDR [90] were used for analyses and data visualizations. The R package tidyverse [91] was used to conduct core analyses (see below and supplementary information 28 for further R packages used).

### Selection of genome-wide association study summary statistics

Genome-wide association study (GWAS) summary statistics were internally generated for three different subunits of the hypothalamus, and externally obtained for ten psychiatric and metabolic conditions (autism, bipolar disorder (BD), blood lipids (high density lipoprotein (HDL) and low density lipoprotein (LDL)), body mass index (BMI), systolic blood pressure (SBP), schizophrenia (SCZ), type II diabetes mellitus (T2D), waist-to-hip ratio (WHR)), to calculate the genetic overlap between selected oxytocinergic/vasotocinergic hypothalamic volumes and these conditions.

To generate hypothalamus GWAS summary statistics, we used the anterior-inferior (aiHyp), anterior-superior (asHyp) and tubular-superior (tsHyp) subunits from the hypothalamus to represent the supraoptic (SON) and paraventricular (PVN) nucleus. The computational process is described in detail below. The aiHyp and asHyp are predominantly comprised of the SON and PVN, respectively, and also include the suprachiasmatic nucleus (containing some vasotocin neurons [17]), and preoptic area [62, 63]. The tsHyp is mainly comprised of large parts of the posterior PVN, with sections of the dorsomedial nucleus and lateral hypothalamus [62, 63]. We decided against including the tubular-inferior (tiHyp) subunit because the subunit seems to be predominantly comprised of other nuclei (i.e., arcuate/infundibular nucleus, ventromedial nucleus, lateral tubular nucleus, tuberomamillary nucleus), whereas the posterior SON appears to only make up for a small fraction of that subunit (compare to [62], [63] also supplementary information, [60]). Thus, we concluded that not including the tiHyp would remove major confounding noise coming from the other nuclei and would not lead to a loss of distinct information from the SON, thus making the analysis more targeted and interpretable. The posterior hypothalamic subunit was not included due to neither containing PVN nor SON material and thus no oxytocin/vasotocin neurons [62].

For autism [92], BD [93] and SCZ [94], the GWAS summary statistics were publicly available from the Psychiatric Genomics Consortium (PGC, https://pgc.unc.edu/). For BMI [95] and WHR [96], the summary statistics are publicly available for download from the GIANT consortium (https://portals.broadinstitute.org/collaboration/giant/index.php/GIANT_consortium). For BMD [97], summary statistics are available at the UK Biobank (UKB, https://www.ukbiobank.ac.uk/. GWAS summary statistics were obtained from the Million Veteran Program (MVP, https://www.research.va.gov/mvp/) via a licensed agreement (dbGaP accession number phs001672) for HDL/LDL [98] and SBP [99], and lastly, they are publicly available from the DIAGRAM consortium (https://diagram-consortium.org/index.html) for T2D [100]. Due to the nature of subsequent conjFDR analyses, ideally, none of the psychiatric/metabolic phenotypes should feature any samples from the UKB. This was true for all phenotypes with the exception of BMD. After exhaustive research for a sufficiently large and recent GWAS and preliminary quality assessments of different obtained summary statistics (e.g., available number of SNPs, missing data), we found that none of the publicly available GWAS summary statistics that both excluded UKB data and consisted of European ancestry were reliable. Therefore, in order to avoid having to exclude BMD from all analyses altogether, we opted for using UKB based summary statistics for the MiXeR analyses and excluding the trait from the subsequent conjFDR analyses. The eventually included GWAS summary statistics for BMD are publicly available from the UKB [97]. Furthermore, the gathered GWAS summary statistics were uniformly based on samples of European/white ancestry. An overview of all included summary statistics is provided in supplementary information 29.

Prior to analyses, all summary statistics underwent preprocessing and cleaning based on the ‘cleansumstats’ pipeline by Gådin and colleagues ([101], https://github.com/BioPsyk/cleansumstats) to harmonize the data frame format. This included, among other things, mapping the input GWAS summary statistics to a dbsnp reference build, standardizing column names and excluding the major histocompatibility complex (MHC) (for details see https://github.com/BioPsyk/cleansumstats/blob/1.6.0/docs/output.md).

### Segmentation of oxytocinergic-vasotocinergic hypothalamus subunits

This study used data from the UK Biobank (UKB), a large-scale biomedical database containing genotype and phenotype data for approximately 500,000 individuals [102]. Prior to participation, all individuals provided written informed consent. The UKB study was approved by the National Health Service National Research Ethics Service (reference: 11/NW/0382). Data for this study were obtained under accession number 27412. The T1-weighted MRI brain images from N = 48,063 genotyped white British individuals were used to segment the hypothalamus into five subunits (anterior-inferior, anterior-superior, tubular-superior, tubular-inferior, posterior) using the automated segmentation tool (version 7.4.1 [62], whole brain segmented with the brain imaging software FreeSurfer version 5.3, [103]). Segmentation did not complete for N = 8 individuals. The segmented data was then prepared for subsequent GWAS, implementing the following processing and quality control steps: i) Regions of interest (ROIs; i.e., five hypothalamus subunits) were averaged across left and right hemisphere, ii) the variables sex, age, age^2^, scanner site, FreeSurfer’s Euler number (a proxy of surface reconstruction and image quality [104]) averaged across hemispheres, total estimated intracranial volume and the first twenty genetic principal components were added, iii) exclusion of subjects with *z*-scores *>* five standard deviations from the mean in any of the structural volume measures for the five ROIs and additionally total estimated intracranial volume, iv) exclusion of individuals *<* 45 and *>* 82 years (mean: 64.89, SD: 7.69), v) exclusion of subjects with extensive movement (Euler score ≤ -200) [104], vi) exclusion of individuals wit nonCaucasian ancestry. This lead to an interim sample of N = 39,712.

### Genome-wide association study on oxytocinergic-vasotocinergic hypothalamus subunits

GWASsummary statistics for the three hypothalamus subunits of interest (anterior-inferior, anterior-superior, and tubular-superior) were generated from version 3 of the UKB genetic data using REGENIE (v3.6 [86]. Genotyping, imputation and central quality control procedures for the UKB genotypes are described in detail elsewhere [105]. We selected white British individuals (as determined by both self-declared ethnicity and principal component analysis) who had undergone the neuroimaging protocol. Participants who had withdrawn their consent were removed, resulting in 39,649 individuals with a mean age of 64.9 years (SD = 7.7 years); 52.1% were female. For the association analysis, we retained only autosomal variants with a minor allele count *>* 20 and an imputation information score *>* 0.8, leaving 16.4 million variants. Sex, age, age^2^, scanner site, FreeSurfer’s Euler number, and the first twenty genetic principal components were included as covariates. Prior to the analysis, volumes of each analyzed hypothalamus subunit were transformed using rank-based inverse normal transformation. The summary statistics were submitted to the “Functional Mapping and Annotation of Genome-Wide Association Studies” (FUMA) web service (version 1.6.3, https://fuma.ctglab.nl, [59]) SNP2GENE in three queries to gather the number of LD-independent loci per subunit GWAS using the default parameter settings in FUMA. Manhattan and QQ plots were directly exported from FUMA.

### Genetic overlap analysis using causal mixture models

Bivariate causal mixture models were fitted for aiHyp, asHyp, tsHyp, autism, BD, SCZ, BMD, BMI, HDL, LDL, SBP, T2D andWHRGWAS summary statistics with the statistical tool MiXeR [55]. Bivariate MiXeR uses Gaussian mixture models to calculate the overall SNP-based genetic overlap, which exists irrespective of genetic correlation, shared between two traits building upon the total number of trait-influencing variants/SNPs linked to each trait individually (considering factors such as LD structure and minor allele frequency). The tool yields a total number of variants and a percentage quantifying the degree of cross-trait overlap. This approach comes with several advantages, such as the discovery of opposite or mixed directions of effect that the same variant may exhibit on two traits (concordant and discordant genetic effects [74]), and the tolerance for sample overlap between two GWAS summary statistics [55]. All causal mixture model analyses were run on an internal secure computing cluster for sensitive data and an adjacent high performance computing cluster using the MiXeR package (version 1.3 [55], https://github.com/precimed/mixer) and bash scripting.

Firstly, in preparation of the bivariate models, univariate models were fitted for each of the thirteen traits separately (aiHyp, asHyp, tsHyp, autism, BD, SCZ, BMD, BMI, HDL, LDL, SBP, T2D, WHR) to estimate the global number of variants associated with each phenotype individually (averaged across 20 independent MiXeR runs). The Akaike information criterion (AIC), where the difference between the AIC of a best-fitting model produced by MiXeR versus an infinitesimal model was compared [74], and heritability (h^2^) estimates using linkage disequilibrium score (LDSC) regression [87] were used to inform the goodness of model fit and reliability of the traits. If the AIC difference was positive and individual *h*^2^ estimates equal to approximately 0.10, we proceeded with bivariate models; if the AIC difference was negative or *h*^2^ estimates for a trait considerably below 0.10, the trait was dropped due to insufficient power. Univariate MiXeR models revealed positive AIC differences and sufficient *h*^2^ estimates for all traits and consequently all were considered for the further analyses (supplementary information 10).

Secondly, expanding from the univariate analysis results, bivariate models were fitted to estimate the global number of trait-influencing variants jointly associated with a phenotype pair. Each combination of hypothalamus subunit and psychiatric/metabolic trait (e.g., aiHyp and autism, aiHyp and T2D, …, asHyp and autism, …, tsHyp and autism, …, etc.) was tested, resulting in a total of 30 combinations or independent models. For the bivariate MiXeR models, two AIC values were used to inform the goodness of model fit, comparing against a (1) minimum and (2) maximum, respectively, possible overlap reference model [74]. A positive difference for one of the two comparisons would indicate good fit. In addition, the log-likelihood distributions for each bivariate model were visually inspected by the first author and co-author AS. For the majority of models, at least one of the two AIC values was *>* 0 and the log-likelihood distribution credible. Both AIC values were negative for aiHyp and autism, and asHyp and autism. For aiHyp and HDL, asHyp and HDL, asHyp and LDL, tsHyp and HDL, and tsHyp and LDL, at least one of the AIC values was positive, however, loglikelihood distribution were not credible. This is indicative of unreliable model fit and non-credible results, which led to the rejection of these seven bivariate models. All relative overlap estimations described in the results were calculated as the proportion of shared variants in the total number of variants estimated for a hypothalamus subunit. As described above, genetic overlap and genetic correlation do not measure the same concept. While the existence of correlation implies existence of genetic overlap, some phenotypes might strongly genetically overlap without substantial correlation. To complement the genetic overlap MiXeR analyses and to compare genetic overlap with genetic correlation, genetic correlations for the same trait pairs were additionally calculated using LDSC regression [89]. Of note, there is no clear consensus on what a ‘small’, ‘medium’ or ‘extensive’ overlap between a pair of traits constitutes in a causal mixture models framework. We therefore draw on related work deploying MiXeR [54, 74, 90] to assess the results.

### Conjunctional false discovery rate for detection of shared genetic loci

To pinpoint the genetic loci underlying the cross-trait genetic overlap previously detected with MiXeR, we leveraged the conjunctional false discovery rate (conjFDR) approach. The conjFDR method uses GWAS summary statistics in a model-free fashion to detect common loci that are associated with two traits at the same time. For this, the method extends the original conditional false discovery rate (condFDR) approach: In a phenotype pair comparison, two separate condFDR analyses for each phenotype using its GWAS summary statistics are conducted with each phenotype as the primary trait conditional on a secondary trait, and vice versa, assigning an FDR value to each SNP, which are then ranked within a Bayesian framework. If the FDR value for a certain SNP remains below a significance threshold (usually *<* 0.05) in both trait condFDRs, the variant will be considered to be associated with the two traits simultaneously [90]. This conjFDR approach increases the chances of discovering novel genetic loci for which conventional single-trait GWAS would be underpowered for and, like the MiXeR method, has the benefit of detecting SNP associations irrespective of genetic correlations while keeping the option of revealing mixed directions of effect *a posteriori*. Here, we performed 20 independent conjFDR analyses for each combination of hypothalamus subunit and psychiatric/metabolic trait (excluding seven unreliable combinations from MiXeR and all combinations featuring BMD), applying a 5% significance threshold, to detect significant loci across traits. Adjacent rsID annotation failed for one locus (chromosome position 44212526).

### Annotation of cross-trait significant loci

To annotate the significant cross-trait variants with general information and surrounding genes, the tool ‘Open Targets Genetics’ (version 22.10 [57, 58], https://genetics.opentargets.org/) was sourced through R using the package ‘otargen’ (version v1.1.1 [106], https://github.com/amirfeizi/otargen). Open Targets Genetics is an analysis pipeline that draws on GWAS data featuring functional genomics data to help interpret e.g., genes or SNPs, and to identify drug targets. Here, we applied the Variant-to-Gene (V2G) score from the ‘genesForVariant’ function (comparable to the ‘locus to gene’ (L2G) score) to fetch all associated genes with meaningful evidence for relevant functional data [107]. The SNPs from each hypothalamus subunit and psychiatric/metabolic trait conjFDR comparison were gathered in one dataset and annotated in bulk, i.e., 124 variants *including* duplicates. Annotation failed for six SNPs (rs647455, rs11598702, rs13332755, rs11191469, rs79353333, rs2293889). The annotated genes were then grouped in six subsets by subunit versus psychiatric or metabolic trait (e.g., aiHyp and psychiatric, aiHyp and metabolic, asHyp and psychiatric, etc.), gene duplicates within each subset were removed. Since there is no consensus V2G threshold provided as significance cut-off [108], we further filtered out less meaningful Open Targets gene annotations by grouping the underlying raw SNPs from the conjFDR analyses into the same six subsets. The following steps were pursued for each SNP subset separately. Within a subset, each SNP was annotated with the closest gene it maps to based on nucleotide distance using the ‘getBM’ function from the biomaRt R suite, leading to a small pool of ‘closest genes’. The V2G scores for this pool were identified and the lowest V2G score was taken as a cut-off or threshold for the genes in the respective subset from the functional gene annotation. All genes with a smaller V2G score than this threshold were discarded. To functionally annotate the genes, they were submitted to FUMA (version 1.6.4 [59], https://fuma.ctglab.nl) GENE2FUNC in six separate queries, one each for the genes associated with aiHyp-and-psychiatric (n_genes_ = 26, aiHyp-and-metabolic (n_genes_ = 125), asHyp-and-psychiatric (n_genes_ = 182), asHyp-and-metabolic (n_genes_ = 67), tsHyp-and-psychiatric (n_genes_ = 70), and tsHyp-and-metabolic (n_genes_ = 151) variants, respectively (analysis settings: background set “Protein-coding”, Ensembl version v110, MHC excluded, default Bonferroni correction for all tests except for gene-set enrichment testing where FDR correction was applied, minimum overlapping genes in gene set annotation set to ≥ 2). The online tool uses GTEx RNA-seq data (version v8, URL, [109]) to test for tissue-specific enrichment of a gene set (here focused on 30 tissues across the human body, including the brain) and performs a hypergeometric test to assess enrichment of genes in different categories (i.e., GWAS) and pathways. For detailed information on data sets and configuration of statistic tests implemented in FUMA, see [59].

## Data Availability

All produced data are available online at https://osf.io/k46gu/

## Acknowledgments

This research was funded by Research Council of Norway (301767). ***Figure 1*** was created with Biorender.com, the grid for the Venn diagrams in ***Figure 3*** was created in Adobe Illustrator 2024.

## Data and code availability

The GWAS summary statistics data is partially publicly available for download at the consortium’s official websites (PGC: https://pgc.unc.edu/, GIANT consortium: https://portals.broadinstitute.org/collaboration/giant/index.php/GIANT_consortium, DIAGRAM consortium: https://diagram-consortium.org/index.html, GEnetic Factors for OSteoporosis Consortium (GEFOS): http://www.gefos.org/?q=content/data-release-2018). Supplementary information, scripts used for analyses with information on specific parameter settings and additional notes are available at https://osf.io/k46gu/.

## Author contributions

AIS, DSQ and DVDM conceived and planned the study. AIS analyzed the data, with contributions from JR for the hypothalamus subunits segmentation, AS for the GWAS, heritability, MiXeR and genetic correlation analyses, and DVDM for the conjFDR analyses. DSQ and DVDM supervised the study. DSQ provided funding for the study. AIS, DSQ, DVDM, AS, JR, MC, AW, OAA, ES, TN and LTW contributed to the interpretation of the results. AIS wrote the first and revised drafts of the manuscript, with DSQ, DVDM, AS, JR, MC, AW, OAA, ES, TN and LTW contributing to the first and revised drafts of the manuscript.

## References

[1] Leng, G., & Leng, R. I. (2021). Oxytocin: A citation network analysis of 10 000 papers. Journal of Neuroendocrinology, 33(11), e13014. 10.1111/jne.13014

[2] Kenkel, W. M. (2019). Corpus colossal: A bibliometric analysis of neuroscience abstracts and impact factor. Frontiers in Integrative Neuroscience, 13. 10.3389/fnint.2019.00018

[3] Howarth, G., & Botha, D. J. (2001). Amniotomy plus intravenous oxytocin for induction of labour. The Cochrane Database of Systematic Reviews, 2001(3), CD003250. 10.1002/14651858.CD003250

[4] Ruis, H., Rolland, R., Doesburg, W., Broeders, G., & Corbey, R. (1981). Oxytocin enhances onset of lactation among mothers delivering prematurely. BMJ, 283(6287), 340–342. 10.1136/bmj.283.6287.340

[5] Bradley, E. R., & Woolley, J. D. (2017). Oxytocin effects in schizophrenia: Reconciling mixed findings and moving forward. Neuroscience & Biobehavioral Reviews, 80, 36–56. 10.1016/j.neubiorev.2017.05.007

[6] Jurek, B., & Neumann, I. D. (2018). The oxytocin receptor: From intracellular signaling to behavior. Physiological Reviews, 98(3), 1805–1908. 10.1152/physrev.00031.2017

[7] Winterton, A., Westlye, L. T., Steen, N. E., Andreassen, O. A., & Quintana, D. S. (2021). Improving the precision of intranasal oxytocin research. Nature Human Behaviour, 5(1), 9–18. 10.1038/s41562-020-00996-4

[8] Vancampfort, D., Stubbs, B., Mitchell, A. J., De Hert, M., Wampers, M., Ward, P. B., Rosenbaum, S., & Correll, C. U. (2015). Risk of metabolic syndrome and its components in people with schizophrenia and related psychotic disorders, bipolar disorder and major depressive disorder: A systematic review and meta-analysis. World Psychiatry, 14(3), 339–347. 10.1002/wps.20252

[9] Birkenaes, A. B., Opjordsmoen, S., Brunborg, C., Engh, J. A., Jonsdottir, H., Ringen, P. A., Simonsen, C., Vaskinn, A., Birkeland, K. I., Friis, S., Sundet, K., & Andreassen, O. A. (2007). The level of cardiovascular risk factors in bipolar disorder equals that of schizophrenia: A comparative study. The Journal of Clinical Psychiatry, 68(6), 10292. 10.4088/JCP.v68n0614

[10] Venkatasubramanian, G., Chittiprol, S., Neelakantachar, N., Naveen, M. N., Thirthall, J., Gangadhar, B. N., & Shetty, K. T. (2007). Insulin and insulin-like growth factor-1 abnormalities in antipsychotic-naive schizophrenia. The American Journal of Psychiatry, 164(10), 1557–1560. 10.1176/appi.ajp.2007.07020233

[11] Winterton, A., Bettella, F., de Lange, A.-M. G., Haram, M., Steen, N. E., Westlye, L. T., Andreassen, O. A., & Quintana, D. S. (2021). Oxytocin-pathway polygenic scores for severe mental disorders and metabolic phenotypes in the UK biobank. Translational Psychiatry, 11(1), 1–9. 10.1038/s41398-021-01725-9

[12] Theofanopoulou, C., Gedman, G., Cahill, J. A., Boeckx, C., & Jarvis, E. D. (2021). Universal nomenclature for oxytocin–vasotocin ligand and receptor families. Nature, 592(7856), 747–755. 10.1038/s41586-020-03040-7

[13] Bernstein, H.-G., Dobrowolny, H., Bogerts, B., Keilhoff, G., & Steiner, J. (2019). The hypothalamus and neuropsychiatric disorders: Psychiatry meets microscopy. Cell and Tissue Research, 375(1), 243–258. 10.1007/s00441-018-2849-3

[14] Kovács, K. J. (2013). CRH: The link between hormonal-, metabolic- and behavioral responses to stress. Journal of Chemical Neuroanatomy, 54, 25–33. 10.1016/j.jchemneu.2013.05.003

[15] Ivell, R. (1986). Biosynthesis of oxytocin in the brain and peripheral organs. In D. Ganten & D. Pfaff (Eds.), Neurobiology of oxytocin (pp. 1–18). Springer. 10.1007/978-3-642-70414-7_1

[16] Leng, G. (2018, July 31). The heart of the brain: The hypothalamus and its hormones. MIT Press.

[17] Buijs, R. M., Soto Tinoco, E. C., Hurtado Alvarado, G., & Escobar, C. (2021, January 1). Chapter 15 - the circadian system: From clocks to physiology. In D. F. Swaab, F. Kreier, P. J. Lucassen, A. Salehi, & R. M. Buijs (Eds.), Handbook of clinical neurology (pp. 233–247, Vol. 179). Elsevier. 10.1016/B978-0-12-819975-6.00013-3

[18] Althammer, F., & Grinevich, V. (2018). Diversity of oxytocin neurones: Beyond magno- and parvocellular cell types? Journal of Neuroendocrinology, 30(8), e12549. 10.1111/jne.12549

[19] Gwee, P.-C., Tay, B.-H., Brenner, S., & Venkatesh, B. (2009). Characterization of the neurohypophysial hormone gene loci in elephant shark and the japanese lamprey: Origin of the vertebrate neurohypophysial hormone genes. BMC Evolutionary Biology, 9(1), 47. 10.1186/1471-2148-9-47

[20] Sartorius, A. M., Rokicki, J., Birkeland, S., Bettella, F., Barth, C., de Lange, A.-M. G., Haram, M., Shadrin, A., Winterton, A., Steen, N. E., Schwarz, E., Stein, D. J., Andreassen, O. A., van der Meer, D., Westlye, L. T., Theofanopoulou, C., & Quintana, D. S. (2024). An evolutionary timeline of the oxytocin signaling pathway. Communications Biology, 7(1), 1–13. 10.1038/s42003-024-06094-9

[21] Gruber, C. W. (2014). Physiology of invertebrate oxytocin and vasopressin neuropeptides. Experimental Physiology, 99(1), 55–61. 10.1113/expphysiol.2013.072561

[22] Richter, D. (1987). Oxytocin and vasopressin genes. In J. F. Habener (Ed.), Molecular cloning of hormone genes (pp. 173–206). Humana Press. 10.1007/978-1-4612-4824-8_8

[23] Bankir, L., Bichet, D. G., & Morgenthaler, N. G. (2017). Vasopressin: Physiology, assessment and osmosensation. Journal of Internal Medicine, 282(4), 284–297. 10.1111/joim.12645

[24] Holt, N. F., & Haspel, K. L. (2010). Vasopressin: A review of therapeutic applications. Journal of Cardiothoracic and Vascular Anesthesia, 24(2), 330–347. 10.1053/j.jvca.2009.09.006

[25] Pasquali, R., Gagliardi, L., Vicennati, V., Gambineri, A., Colitta, D., Ceroni, L., & Casimirri, F. (1999). ACTH and cortisol response to combined corticotropin releasing hormone-arginine vasopressin stimulation in obese males and its relationship to body weight, fat distribution and parameters of the metabolic syndrome. International Journal of Obesity, 23(4), 419–424. 10.1038/sj.ijo.0800838

[26] Heinrichs, M., von Dawans, B., & Domes, G. (2009). Oxytocin, vasopressin, and human social behavior. Frontiers in Neuroendocrinology, 30(4), 548–557. 10.1016/j.yfrne.2009.05.005

[27] Keverne, E. B., & Curley, J. P. (2004). Vasopressin, oxytocin and social behaviour. Current Opinion in Neurobiology, 14(6), 777–783. 10.1016/j.conb.2004.10.006

[28] Song, Z., & Albers, H. E. (2018). Cross-talk among oxytocin and arginine-vasopressin receptors: Relevance for basic and clinical studies of the brain and periphery. Frontiers in Neuroendocrinology, 51, 14–24. 10.1016/j.yfrne.2017.10.004

[29] Kreier, F., & Swaab, D. F. (2021, January 1). Chapter 2 - history of hypothalamic research: “the spring of primitive existence”;. In D. F. Swaab, F. Kreier, P. J. Lucassen, A. Salehi, & R. M. Buijs (Eds.), Handbook of clinical neurology (pp. 7–43, Vol. 179). Elsevier. 10.1016/B978-0-12-819975-6.00031-5

[30] Leibowitz, S. F., & Alexander, J. T. (1998). Hypothalamic serotonin in control of eating behavior, meal size, and body weight. Biological Psychiatry, 44(9), 851–864. 10.1016/S0006-3223(98)00186-3

[31] Saper, C. B., Chou, T. C., & Elmquist, J. K. (2002). The need to feed: Homeostatic and hedonic control of eating. Neuron, 36(2), 199–211. 10.1016/S0896-6273(02)00969-8

[32] Saper, C. B., Scammell, T. E., & Lu, J. (2005). Hypothalamic regulation of sleep and circadian rhythms. Nature, 437(7063), 1257–1263. 10.1038/nature04284

[33] Le, T. M., Liao, D.-L., Ide, J., Zhang, S., Zhornitsky, S., Wang, W., & Li, C.-S. R. (2020). The interrelationship of body mass index with gray matter volume and resting-state functional connectivity of the hypothalamus. International Journal of Obesity, 44(5), 1097–1107. 10.1038/s41366-019-0496-8

[34] Brown, S. S. G., Westwater, M. L., Seidlitz, J., Ziauddeen, H., & Fletcher, P. C. (2023). Hypothalamic volume is associated with body mass index. NeuroImage: Clinical, 39, 103478. 10.1016/j.nicl.2023.103478

[35] Rahmouni, K. (2016). Cardiovascular regulation by the arcuate nucleus of the hypothalamus. Hypertension, 67(6), 1064–1071. 10.1161/HYPERTENSIONAHA.115.06425

[36] Sapru, H. N. (2013). Role of the hypothalamic arcuate nucleus in cardiovascular regulation. Autonomic Neuroscience: Basic and Clinical, 175(1), 38–50. 10.1016/j.autneu.2012.10.016

[37] Guillemin, R., & Burgus, R. (1972). The hormones of the hypothalamus. Scientific American, 227(5), 24–33. 10.1038/scientificamerican1172-24

[38] Bernstein, H.-G., Müller, S., Dobrowolny, H., Wolke, C., Lendeckel, U., Bukowska, A., Keilhoff, G., Becker, A., Trübner, K., Steiner, J., & Bogerts, B. (2017). Insulin-regulated aminopeptidase immunoreactivity is abundantly present in human hypothalamus and posterior pituitary gland, with reduced expression in paraventricular and suprachiasmatic neurons in chronic schizophrenia. European Archives of Psychiatry and Clinical Neuroscience, 267(5), 427–443. 10.1007/s00406-016-0757-7

[39] Guo, L., Qi, Y.-J., Tan, H., Dai, D., Balesar, R., Sluiter, A., Heerikhuize, J. v., Hu, S.-H., Swaab, D. F., & Bao, A.-M. (2022). Different oxytocin and corticotropin-releasing hormone system changes in bipolar disorder and major depressive disorder patients. eBioMedicine, 84. 10.1016/j.ebiom.2022.104266

[40] Ross, H. E., & Young, L. J. (2009). Oxytocin and the neural mechanisms regulating social cognition and affiliative behavior. Frontiers in Neuroendocrinology, 30(4), 534–547. 10.1016/j.yfrne.2009.05.004

[41] Inada, K., Tsujimoto, K., Yoshida, M., Nishimori, K., & Miyamichi, K. (2022). Oxytocin signaling in the posterior hypothalamus prevents hyperphagic obesity in mice (J. K. Elmquist & M.-L. Wong, Eds.). eLife, 11, e75718. 10.7554/eLife.75718

[42] Chen, S.-D., You, J., Zhang, W., Wu, B.-S., Ge, Y.-J., Xiang, S.-T., Du, J., Kuo, K., Banaschewski, T., Barker, G. J., Bokde, A. L. W., Desrivières, S., Flor, H., Grigis, A., Garavan, H., Gowland, P., Heinz, A., Brühl, R., Martinot, J.-L., … Yu, J.-T. (2024). The genetic architecture of the human hypothalamus and its involvement in neuropsychiatric behaviours and disorders. Nature Human Behaviour, 1–15. 10.1038/s41562-023-01792-6

[43] Satizabal, C. L., Adams, H. H. H., Hibar, D. P., White, C. C., Knol, M. J., Stein, J. L., Scholz, M., Sargurupremraj, M., Jahanshad, N., Roshchupkin, G. V., Smith, A. V., Bis, J. C., Jian, X., Luciano, M., Hofer, E., Teumer, A., van der Lee, S. J., Yang, J., Yanek, L. R., … Ikram, M. A. (2019). Genetic architecture of subcortical brain structures in 38,851 individuals. Nature Genetics, 51(11), 1624–1636. 10.1038/s41588-019-0511-y

[44] Vattikuti, S., Guo, J., & Chow, C. C. (2012). Heritability and genetic correlations explained by common SNPs for metabolic syndrome traits. PLOS Genetics, 8(3), e1002637. 10.1371/journal.pgen.1002637

[45] DeForest, N., & Majithia, A. R. (2022). Genetics of type 2 diabetes: Implications from large-scale studies. Current Diabetes Reports, 22(5), 227–235. 10.1007/s11892-022-01462-3

[46] Hilker, R., Helenius, D., Fagerlund, B., Skytthe, A., Christensen, K., Werge, T. M., Nordentoft, M., & Glenthøj, B. (2018). Heritability of schizophrenia and schizophrenia spectrum based on the nationwide danish twin register. Biological Psychiatry, 83(6), 492–498. 10.1016/j.biopsych.2017.08.017

[47] Gordovez, F. J. A., & McMahon, F. J. (2020). The genetics of bipolar disorder. Molecular Psychiatry, 25(3), 544–559. 10.1038/s41380-019-0634-7

[48] Hibar, D. P., Stein, J. L., Renteria, M. E., Arias-Vasquez, A., Desrivières, S., Jahanshad, N., Toro, R., Wittfeld, K., Abramovic, L., Andersson, M., Aribisala, B. S., Armstrong, N. J., Bernard, M., Bohlken, M. M., Boks, M. P., Bralten, J., Brown, A. A., Mallar Chakravarty, M., Chen, Q., … Medland, S. E. (2015). Common genetic variants influence human subcortical brain structures. Nature, 520(7546), 224–229. 10.1038/nature14101

[49] Hibar, D. P., Adams, H. H. H., Jahanshad, N., Chauhan, G., Stein, J. L., Hofer, E., Renteria, M. E., Bis, J. C., Arias-Vasquez, A., Ikram, M. K., Desrivières, S., Vernooij, M. W., Abramovic, L., Alhusaini, S., Amin, N., Andersson, M., Arfanakis, K., Aribisala, B. S., Armstrong, N. J., … Ikram, M. A. (2017). Novel genetic loci associated with hippocampal volume. Nature Communications, 8(1), 13624. 10.1038/ncomms13624

[50] Elvsåshagen, T., Shadrin, A., Frei, O., van der Meer, D., Bahrami, S., Kumar, V. J., Smeland, O., Westlye, L. T., Andreassen, O. A., & Kaufmann, T. (2021). The genetic architecture of the human thalamus and its overlap with ten common brain disorders. Nature Communications, 12(1), 2909. 10.1038/s41467-021-23175-z

[51] Elvsåshagen, T., Bahrami, S., van der Meer, D., Agartz, I., Alnæs, D., Barch, D. M., Baur-Streubel, R., Bertolino, A., Beyer, M. K., Blasi, G., Borgwardt, S., Boye, B., Buitelaar, J., Bøen, E., Celius, E. G., Cervenka, S., Conzelmann, A., Coynel, D., Di Carlo, P., … Kaufmann, T. (2020). The genetic architecture of human brainstem structures and their involvement in common brain disorders. Nature Communications, 11(1), 4016. 10.1038/s41467-020-17376-1

[52] Bahrami, S., Nordengen, K., Rokicki, J., Shadrin, A. A., Rahman, Z., Smeland, O. B., Jaholkowski, P. P., Parker, N., Parekh, P., O’Connell, K. S., Elvsåshagen, T., Toft, M., Djurovic, S., Dale, A. M., Westlye, L. T., Kaufmann, T., & Andreassen, O. A. (2024). The genetic landscape of basal ganglia and implications for common brain disorders. Nature Communications, 15(1), 8476. 10.1038/s41467-024-52583-0

[53] Moberget, T., Meer, D. v. d., Bahrami, S., Roelfs, D., Frei, O., Kaufmann, T., Fernandez-Cabello, S., Kim, M., Wolfers, T., Diedrichsen, J., Smeland, O. B., Shadrin, A., Dale, A., Andreassen, O. A., & Westlye, L. T. (2024, January 24). The genetic architecture of human cerebellar morphology supports a key role for the cerebellum in human evolution and psychopathology. 10.1101/2023.02.10.23285704

[54] Campbell, M. L., Shadrin, A., Meer, D. v. d., Tesfaye, M., Smeland, O. B., O’Connell, K. S., Parker, N., Frei, O., Andreassen, O., Stein, D. J., Dalvie, S., & Rokicki, J. (2023, December 4). Regional genetic architecture of the corpus callosum and its overlap with psychiatric and substance use phenotypes. 10.1101/2023.12.03.23299351

[55] Frei, O., Holland, D., Smeland, O. B., Shadrin, A. A., Fan, C. C., Maeland, S., O’Connell, K. S., Wang, Y., Djurovic, S., Thompson, W. K., Andreassen, O. A., & Dale, A. M. (2019). Bivariate causal mixture model quantifies polygenic overlap between complex traits beyond genetic correlation. Nature Communications, 10(1), 2417. 10.1038/s41467-019-10310-0

[56] Andreassen, O. A., Thompson, W. K., Schork, A. J., Ripke, S., Mattingsdal, M., Kelsoe, J. R., Kendler, K. S., O’Donovan, M. C., Rujescu, D., Werge, T., Sklar, P., Consortium (PGC), T. P. G., Groups, B. D. bibinitperiod S. W., Roddey, J. C., Chen, C.-H., McEvoy, L., Desikan, R. S., Djurovic, S., & Dale, A. M. (2013). Improved detection of common variants associated with schizophrenia and bipolar disorder using pleiotropy-informed conditional false discovery rate. PLOS Genetics, 9(4), e1003455. 10.1371/journal.pgen.1003455

[57] Mountjoy, E., Schmidt, E. M., Carmona, M., Schwartzentruber, J., Peat, G., Miranda, A., Fumis, L., Hayhurst, J., Buniello, A., Karim, M. A., Wright, D., Hercules, A., Papa, E., Fauman, E. B., Barrett, J. C., Todd, J. A., Ochoa, D., Dunham, I., & Ghoussaini, M. (2021). An open approach to systematically prioritize causal variants and genes at all published human GWAS trait-associated loci. Nature Genetics, 53(11), 1527–1533. 10.1038/s41588-021-00945-5

[58] Ghoussaini, M., Mountjoy, E., Carmona, M., Peat, G., Schmidt, E. M., Hercules, A., Fumis, L., Miranda, A., Carvalho-Silva, D., Buniello, A., Burdett, T., Hayhurst, J., Baker, J., Ferrer, J., Gonzalez-Uriarte, A., Jupp, S., Karim, M. A., Koscielny, G., Machlitt-Northen, S., … Dunham, I. (2021). Open targets genetics: Systematic identification of trait-associated genes using large-scale genetics and functional genomics. Nucleic Acids Research, 49, D1311–D1320. 10.1093/nar/gkaa840

[59] Watanabe, K., Taskesen, E., van Bochoven, A., & Posthuma, D. (2017). Functional mapping and annotation of genetic associations with FUMA. Nature Communications, 8(1), 1826. 10.1038/s41467-017-01261-5

[60] Shapiro, N. L., Todd, E. G., Billot, B., Cash, D. M., Iglesias, J. E., Warren, J. D., Rohrer, J. D., & Bocchetta, M. (2022). In vivo hypothalamic regional volumetry across the frontotemporal dementia spectrum. NeuroImage: Clinical, 35, 103084. 10.1016/j.nicl.2022.103084

[61] Ishunina, T. A., & Swaab, D. F. (1999). Vasopressin and oxytocin neurons of the human supraoptic and paraventricular nucleus; size changes in relation to age and sex. The Journal of Clinical Endocrinology & Metabolism, 84(12), 4637–4644. https://doi.org/2016092613301700987

[62] Billot, B., Bocchetta, M., Todd, E., Dalca, A. V., Rohrer, J. D., & Iglesias, J. E. (2020). Automated segmentation of the hypothalamus and associated subunits in brain MRI. NeuroImage, 223, 117287. 10.1016/j.neuroimage.2020.117287

[63] Bocchetta, M., Gordon, E., Manning, E., Barnes, J., Cash, D. M., Espak, M., Thomas, D. L., Modat, M., Rossor, M. N., Warren, J. D., Ourselin, S., Frisoni, G. B., & Rohrer, J. D. (2015). Detailed volumetric analysis of the hypothalamus in behavioral variant frontotemporal dementia. Journal of Neurology, 262(12), 2635–2642. 10.1007/s00415-015-7885-2

[64] Winterton, A., Bettella, F., Beck, D., Gurholt, T. P., Steen, N. E., Rødevand, L., Westlye, L. T., Andreassen, O. A., & Quintana, D. S. (2022). The oxytocin signalling gene pathway contributes to the association between loneliness and cardiometabolic health. Psychoneuroendocrinology, 144, 105875. 10.1016/j.psyneuen.2022.105875

[65] O’Connor, L. J., Schoech, A. P., Hormozdiari, F., Gazal, S., Patterson, N., & Price, A. L. (2019). Extreme polygenicity of complex traits is explained by negative selection. American Journal of Human Genetics, 105(3), 456–476. 10.1016/j.ajhg.2019.07.003

[66] Lappalainen, T., Li, Y. I., Ramachandran, S., & Gusev, A. (2024). Genetic and molecular architecture of complex traits. Cell, 187(5), 1059–1075. 10.1016/j.cell.2024.01.023

[67] Wendt, F. R., Pathak, G. A., Tylee, D. S., Goswami, A., & Polimanti, R. (2020). Heterogeneity and polygenicity in psychiatric disorders: A genome-wide perspective. Chronic Stress, 4, 2470547020924844. 10.1177/2470547020924844

[68] Doernberg, E., & Hollander, E. (2016). Neurodevelopmental disorders (ASD and ADHD): DSM-5, ICD-10, and ICD-11. CNS Spectrums, 21(4), 295–299. 10.1017/S1092852916000262

[69] Grzadzinski, R., Huerta, M., & Lord, C. (2013). DSM-5 and autism spectrum disorders (ASDs): An opportunity for identifying ASD subtypes. Molecular autism, 4(1). 10.1186/2040-2392-4-12

[70] Greenwood, T. A., Shutes-David, A., & Tsuang, D. W. (2019). Endophenotypes in schizophrenia: Digging deeper to identify genetic mechanisms. Journal of psychiatry and brain science, 4(2), e190005. 10.20900/jpbs.20190005

[71] Donati, F. L., D’Agostino, A., & Ferrarelli, F. (2020). Neurocognitive and neurophysiological endophenotypes in schizophrenia: An overview. Biomarkers in Neuropsychiatry, 3, 100017. 10.1016/j.bionps.2020.100017

[72] Gazal, S., Finucane, H. K., Furlotte, N. A., Loh, P.-R., Palamara, P. F., Liu, X., Schoech, A., Bulik-Sullivan, B., Neale, B. M., Gusev, A., & Price, A. L. (2017). Linkage disequilibrium–dependent architecture of human complex traits shows action of negative selection. Nature Genetics, 49(10), 1421–1427. 10.1038/ng.3954

[73] Boyle, E. A., Li, Y. I., & Pritchard, J. K. (2017). An expanded view of complex traits: From polygenic to omnigenic. Cell, 169(7), 1177–1186. 10.1016/j.cell.2017.05.038

[74] Hindley, G., Frei, O., Shadrin, A. A., Cheng, W., O’Connell, K. S., Icick, R., Parker, N., Bahrami, S., Karadag, N., Roelfs, D., Holen, B., Lin, A., Fan, C. C., Djurovic, S., Dale, A. M., Smeland, O. B., & Andreassen, O. A. (2022). Charting the landscape of genetic overlap between mental disorders and related traits beyond genetic correlation. The American Journal of Psychiatry, 179(11), 833–843. 10.1176/appi.ajp.21101051

[75] Nogueira, F. N., & Carvalho, R. A. (2017). Metabolic remodeling triggered by salivation and diabetes in major salivary glands. NMR in Biomedicine, 30(2), e3683. 10.1002/nbm.3683 e3683 NBM-16-0087.R2.

[76] Rui, L. (2014). Energy metabolism in the liver. Comprehensive Physiology, 4(1), 177–197. 10.1002/cphy.c130024

[77] Lambert, J.-C., Ibrahim-Verbaas, C. A., Harold, D., Naj, A. C., Sims, R., Bellenguez, C., Jun, G., DeStefano, A. L., Bis, J. C., Beecham, G. W., Grenier-Boley, B., Russo, G., Thornton-Wells, T. A., Jones, N., Smith, A. V., Chouraki, V., Thomas, C., Ikram, M. A., Zelenika, D., … Amouyel, P. (2013). Meta-analysis of 74,046 individuals identifies 11 new susceptibility loci for alzheimer’s disease. Nature Genetics, 45(12), 1452–1458. 10.1038/ng.2802

[78] Nho, K., Kim, S., Horgusluoglu, E., Risacher, S. L., Shen, L., Kim, D., Lee, S., Foroud, T., Shaw, L. M., Trojanowski, J. Q., Aisen, P. S., Petersen, R. C., Jack, C. R., Weiner, M. W., Green, R. C., Toga, A. W., Saykin, A. J., & for the Alzheimer’s Disease Neuroimaging Initiative (ADNI). (2017). Association analysis of rare variants near the APOE region with CSF and neuroimaging biomarkers of alzheimer’s disease. BMC Medical Genomics, 10(1), 29. 10.1186/s12920-017-0267-0

[79] Kulminski, A. M., Philipp, I., Shu, L., & Culminskaya, I. (2022). Definitive roles of TOMM40-APOE-APOC1 variants in the alzheimer’s risk. Neurobiology of Aging, 110, 122–131. 10.1016/j.neurobiolaging.2021.09.009

[80] Fekete, C., Légrádi, G., Mihály, E., Huang, Q.-H., Tatro, J. B., Rand, W. M., Emerson, C. H., & Lechan, R. M. (2000). A-melanocyte-stimulating hormone is vontained in nerve terminals innervating thyrotropin-releasing hormone-synthesizing neurons in the hypothalamic paraventricular nucleus and prevents fasting-induced suppression of prothyrotropin-releasing hormone gene expression. Journal of Neuroscience, 20(4), 1550–1558. 10.1523/JNEUROSCI.20-04-01550.2000

[81] Jeong, J. H., Lee, D. K., & Jo, Y.-H. (2017). Cholinergic neurons in the dorsomedial hypothalamus regulate food intake. Molecular Metabolism, 6(3), 306–312. 10.1016/j.molmet.2017.01.001

[82] Saper, C. B., & Lowell, B. B. (2014). The hypothalamus. Current Biology, 24(23), R1111–R1116. 10.1016/j.cub.2014.10.023

[83] Border, R., O’Rourke, S., de Candia, T., Goddard, M. E., Visscher, P. M., Yengo, L., Jones, M., & Keller, M. C. (2022). Assortative mating biases marker-based heritability estimators. Nature Communications, 13(1), 660. 10.1038/s41467-022-28294-9

[84] R Core Team. (2022). R: A language and environment for statistical computing. R Foundation for Statistical Computing. Vienna, Austria. https://www.R-project.org/

[85] Posit team. (2023). Rstudio: Integrated development environment for r. RStudio, PBC. Boston, MA. http://www.rstudio.com/

[86] Mbatchou, J., Barnard, L., Backman, J., Marcketta, A., Kosmicki, J. A., Ziyatdinov, A., Benner, C., O’Dushlaine, C., Barber, M., Boutkov, B., Habegger, L., Ferreira, M., Baras, A., Reid, J., Abecasis, G., Maxwell, E., & Marchini, J. (2021). Computationally efficient whole-genome regression for quantitative and binary traits. Nature Genetics, 53(7), 1097–1103. 10.1038/s41588-021-00870-7

[87] Bulik-Sullivan, B. K., Loh, P.-R., Finucane, H. K., Ripke, S., Yang, J., Patterson, N., Daly, M. J., Price, A. L., & Neale, B. M. (2015). LD score regression distinguishes confounding from polygenicity in genome-wide association studies. Nature Genetics, 47(3), 291–295. 10.1038/ng.3211

[88] Finucane, H. K., Bulik-Sullivan, B., Gusev, A., Trynka, G., Reshef, Y., Loh, P.-R., Anttila, V., Xu, H., Zang, C., Farh, K., Ripke, S., Day, F. R., Purcell, S., Stahl, E., Lindstrom, S., Perry, J. R. B., Okada, Y., Raychaudhuri, S., Daly, M. J., … Price, A. L. (2015). Partitioning heritability by functional annotation using genome-wide association summary statistics. Nature Genetics, 47(11), 1228–1235. 10.1038/ng.3404

[89] Bulik-Sullivan, B., Finucane, H. K., Anttila, V., Gusev, A., Day, F. R., Loh, P.-R., Duncan, L., Perry, J. R. B., Patterson, N., Robinson, E. B., Daly, M. J., Price, A. L., & Neale, B. M. (2015). An atlas of genetic correlations across human diseases and traits. Nature Genetics, 47(11), 1236–1241. 10.1038/ng.3406

[90] Smeland, O. B., Frei, O., Shadrin, A., O’Connell, K., Fan, C.-C., Bahrami, S., Holland, D., Djurovic, S., Thompson, W. K., Dale, A. M., & Andreassen, O. A. (2020). Discovery of shared genomic loci using the conditional false discovery rate approach. Human Genetics, 139(1), 85–94. 10.1007/s00439-019-02060-2

[91] Wickham, H., Averick, M., Bryan, J., Chang, W., McGowan, L. D., François, R., Grolemund, G., Hayes, A., Henry, L., Hester, J., Kuhn, M., Pedersen, T. L., Miller, E., Bache, S. M., Müller, K., Ooms, J., Robinson, D., Seidel, D. P., Spinu, V., … Yutani, H. (2019). Welcome to the tidyverse. Journal of Open Source Software, 4(43), 1686. 10.21105/joss.01686

[92] Grove, J., Ripke, S., Als, T. D., Mattheisen, M., Walters, R. K., Won, H., Pallesen, J., Agerbo, E., Andreassen, O. A., Anney, R., Awashti, S., Belliveau, R., Bettella, F., Buxbaum, J. D., Bybjerg-Grauholm, J., Bækvad-Hansen, M., Cerrato, F., Chambert, K., Christensen, J. H., … Børglum, A. D. (2019). Identification of common genetic risk variants for autism spectrum disorder. Nature Genetics, 51(3), 431–444. 10.1038/s41588-019-0344-8

[93] O’Connell, K. S., Koromina, M., Veen, T. v. d., Boltz, T., David, F. S., Yang, J. M. K., Lin, K.-H., Wang, X., Coleman, J. R. I., Mitchell, B. L., McGrouther, C. C., Rangan, A. V., Lind, P. A., Koch, E., Harder, A., Parker, N., Bendl, J., Adorjan, K., Agerbo, E., … Consortium, t. B. D. W. G. o. t. P. G. (2024, August 28). Genomics yields biological and phenotypic insights into bipolar disorder. 10.1101/2023.10.07.23296687

[94] Trubetskoy, V., Pardiñas, A. F., Qi, T., Panagiotaropoulou, G., Awasthi, S., Bigdeli, T. B., Bryois, J., Chen, C.-Y., Dennison, C. A., Hall, L. S., Lam, M., Watanabe, K., Frei, O., Ge, T., Harwood, J. C., Koopmans, F., Magnusson, S., Richards, A. L., Sidorenko, J., … O’Donovan, M. C. (2022). Mapping genomic loci implicates genes and synaptic biology in schizophrenia. Nature, 604(7906), 502–508. 10.1038/s41586-022-04434-5

[95] Locke, A. E., Kahali, B., Berndt, S. I., Justice, A. E., Pers, T. H., Day, F. R., Powell, C., Vedantam, S., Buchkovich, M. L., Yang, J., Croteau-Chonka, D. C., Esko, T., Fall, T., Ferreira, T., Gustafsson, S., Kutalik, Z., Luan, J., Mägi, R., Randall, J. C., … Speliotes, E. K. (2015). Genetic studies of body mass index yield new insights for obesity biology. Nature, 518(7538), 197–206. 10.1038/nature14177

[96] Shungin, D., Winkler, T. W., Croteau-Chonka, D. C., Ferreira, T., Locke, A. E., Mägi, R., Strawbridge, R. J., Pers, T. H., Fischer, K., Justice, A. E., Workalemahu, T., Wu, J. M. W., Buchkovich, M. L., Heard-Costa, N. L., Roman, T. S., Drong, A. W., Song, C., Gustafsson, S., Day, F. R., … Mohlke, K. L. (2015). New genetic loci link adipose and insulin biology to body fat distribution. Nature, 518(7538), 187–196. 10.1038/nature14132

[97] Morris, J. A., Kemp, J. P., Youlten, S. E., Laurent, L., Logan, J. G., Chai, R. C., Vulpescu, N. A., Forgetta, V., Kleinman, A., Mohanty, S. T., Sergio, C. M., Quinn, J., Nguyen-Yamamoto, L., Luco, A.-L., Vijay, J., Simon, M.-M., Pramatarova, A., Medina-Gomez, C., Trajanoska, K., … Richards, J. B. (2019). An atlas of genetic influences on osteoporosis in humans and mice. Nature Genetics, 51(2), 258–266. 10.1038/s41588-018-0302-x

[98] Klarin, D., Damrauer, S. M., Cho, K., Sun, Y. V., Teslovich, T. M., Honerlaw, J., Gagnon, D. R., DuVall, S. L., Li, J., Peloso, G. M., Chaffin, M., Small, A. M., Huang, J., Tang, H., Lynch, J. A., Ho, Y.-L., Liu, D. J., Emdin, C. A., Li, A. H., … Assimes, T. L. (2018). Genetics of blood lipids among ∼300,000 multi-ethnic participants of the million veteran program. Nature Genetics, 50(11), 1514–1523. 10.1038/s41588-018-0222-9

[99] Giri, A., Hellwege, J. N., Keaton, J. M., Park, J., Qiu, C., Warren, H. R., Torstenson, E. S., Kovesdy, C. P., Sun, Y. V., Wilson, O. D., Robinson-Cohen, C., Roumie, C. L., Chung, C. P., Birdwell, K. A., Damrauer, S. M., DuVall, S. L., Klarin, D., Cho, K., Wang, Y., … Edwards, T. L. (2019). Trans-ethnic association study of blood pressure determinants in over 750,000 individuals. Nature Genetics, 51(1), 51–62. 10.1038/s41588-018-0303-9

[100] Replication, T. D. G., Consortium, M.-a., Morris, A. P., Voight, B. F., Teslovich, T. M., Ferreira, T., Segrè, A. V., Steinthorsdottir, V., Strawbridge, R. J., Khan, H., Grallert, H., Mahajan, A., Prokopenko, I., Kang, H. M., Dina, C., Esko, T., Fraser, R. M., Kanoni, S., Kumar, A., … South Asian Type 2 Diabetes (SAT2D) Consortium. (2012). Large-scale association analysis provides insights into the genetic architecture and pathophysiology of type 2 diabetes. Nature Genetics, 44(9), 981–990. 10.1038/ng.2383

[101] Gadin, J. R., Zetterberg, R., Meijsen, J., & Schork, A. J. (2022, December 15). Cleansumstats: Converting GWAS sumstats to a common format to facilitate downstream applications (Version 1.5.4). Zenodo. 10.5281/zenodo.7540572

[102] Sudlow, C., Gallacher, J., Allen, N., Beral, V., Burton, P., Danesh, J., Downey, P., Elliott, P., Green, J., Landray, M., Liu, B., Matthews, P., Ong, G., Pell, J., Silman, A., Young, A., Sprosen, T., Peakman, T., & Collins, R. (2015). UK biobank: An open access resource for identifying the causes of a wide range of complex diseases of middle and old age. PLOS Medicine, 12(3), e1001779. 10.1371/journal.pmed.1001779

[103] Fischl, B. (2012). FreeSurfer. NeuroImage, 62(2), 774–781. 10.1016/j.neuroimage.2012.01.021

[104] Rosen, A. F. G., Roalf, D. R., Ruparel, K., Blake, J., Seelaus, K., Villa, L. P., Ciric, R., Cook, P. A., Davatzikos, C., Elliott, M. A., Garcia de La Garza, A., Gennatas, E. D., Quarmley, M., Schmitt, J. E., Shinohara, R. T., Tisdall, M. D., Craddock, R. C., Gur, R. E., Gur, R. C., & Satterthwaite, T. D. (2018). Quantitative assessment of structural image quality. NeuroImage, 169, 407–418. 10.1016/j.neuroimage.2017.12.059

[105] Bycroft, C., Freeman, C., Petkova, D., Band, G., Elliott, L. T., Sharp, K., Motyer, A., Vukcevic, D., Delaneau, O., O’Connell, J., Cortes, A., Welsh, S., Young, A., Effingham, M., McVean, G., Leslie, S., Allen, N., Donnelly, P., & Marchini, J. (2018). The UK biobank resource with deep phenotyping and genomic data. Nature, 562(7726), 203–209. 10.1038/s41586-018-0579-z

[106] Feizi, A., & Ray, K. (2023). Otargen: GraphQL-based r package for tidy data accessing and processing from open targets genetics. Bioinformatics, 39(8), btad441. 10.1093/bioinformatics/btad441

[107] Genetics, O. T. (2022, August 16). Assigning variants to genes (v2g) | open targets genetics documentation. https://genetics-docs.opentargets.org/our-approach/data-pipeline

[108] Genetics, O. T. (2023, April 12). FAQs | open targets genetics documentation. https://genetics-docs.opentargets.org/faqs

[109] GTEx-Consortium. (2015). The genotype-tissue expression (GTEx) pilot analysis: Multitissue gene regulation in humans. Science, 348(6235), 648–660. 10.1126/science.1262110

